# Dose-response relationship among body mass index, abdominal adiposity and atrial fibrillation in patients undergoing cardiac surgery: a meta-analysis of 30 prospective studies

**DOI:** 10.1101/2020.10.15.20213595

**Authors:** Menglu Liu, Kaibo Mei, Jianyong Ma, Peng Yu, Lixia Xie, Yujie Zhao, Xiao Liu

**Affiliations:** Department of Cardiology, The Seventh People’s Hospital of Zhengzhou, Zhengzhou, Henan, 450016, China; Anesthesiology Department, the People’s Hospital of Shanggrao, Jiangxi, 330006, China; Department of Pharmacology and Systems Physiology University of Cincinnati College of Medicine, Cincinnati, OH 45267; Department of Endocrine, the Second Affiliated Hospital of Nanchang University, Jiangxi, 330006, China; Department of Respiratory and Critical Care Medicine, the Second Affiliated Hospital of Nanchang University, Jiangxi, 330006, China; Department of Cardiology, the Second Affiliated Hospital of Nanchang University, Jiangxi, 330006, China

**Keywords:** obesity paradox, atrial fibrillation, dose-response, meta-analysis, body mass index, risk factor, overweight, obesity, waist circumference, cardiac surgery, coronary artery bypass graft

## Abstract

**Background:** Whether being overweight increases the risk of postoperative atrial fibrillation (POAF) is unclear, and whether adiposity independently contributes to POAF has not been comprehensively studied. Thus, we conducted a meta-analysis to clarify the strength and shape of the exposure-effect relationship between adiposity and POAF.

**Methods:** The PubMed, Cochrane Library, and EMBASE databases were searched for prospective studies (RCTs, cohort studies, and nest-case control studies) reporting data regarding the relationship between adiposity and the risk of POAF.

**Results:** Thirty publications involving 139,302 patients were included. Analysis of categorical variables showed that obesity (RR: 1.39, P<0.001), but not being underweight (RR: 1.44, P=0.13) or being overweight (RR: 1.03, P=0.48), was associated with an increased risk of POAF. In the exposure-effect analysis, the summary RR for a 5-unit increment in body mass index (BMI) was 1.09 (P<0.001) for the risk of POAF. There was a significant linear relationship between BMI and POAF (P_nonlinearity_=0.91); the curve was flat and began to rise steeply at a BMI of approximately 30. Notably, BMI levels below 30 (overweight) were not associated with a higher risk of POAF. In the subgroup analysis of surgery types, the pooled RR values for a BMI increase of 5 for coronary artery bypass graft and valve surgery were 1.21 (P<0.01) and 1.34 (P=0.25), respectively, suggesting that a potential difference in the association exists by surgery type. Additionally, waist obesity was associated with the risk of POAF (RR: 1.55, P<0.001).

**Conclusion:** Based on the current evidence, our findings show that adiposity was independently associated with an increased risk of POAF, while being underweight or overweight might not significantly increase the POAF risk. The magnitude of the effect of obesity on AF in patients undergoing valve surgery might be small, and this finding needs to be further confirmed.

## Introduction

The prevalence of overweight and obesity has rapidly increased in recent decades in both developing and developed counties^[1-3]^. This increase has raised serious public health concerns due to the positive association between overweight and obesity and an increased risk of various chronic diseases, including cardiovascular diseases, metabolic syndrome, all-cause mortality, and several types of cancer^[4-7]^.

Postoperative atrial fibrillation (POAF) is among the most common complications arising from cardiac surgery, affecting between 20% and 50% of patients, and is associated with significantly worse adverse outcomes (such as all-cause death and stroke)^[8]^. Several prospective studies have reported increased POAF with a higher body mass index^[9-11]^ (BMI, kg/m^2^). Although various studies reported a positive association between obesity (BMI>30) and POAF in patients undergoing cardiac surgery, whether being overweight (BMI of 25-29.9) increases the risk remains unclear as some studies found a positive association^[12, 13]^, while other studies did not^[14-16]^. Several well-designed meta-analyses based on a categorical or continuous model found an increased risk of POAF among individuals with obesity. These studies provide valuable information. However, these studies have several limitations. First, it is arguable that the use of a categorical model in meta-analyses has the risk of reducing power and precision by dividing the exposure into several groups^[17]^. Furthermore, the use of a continuous model might not detect the dose-specific association as several studies have also reported a “U”-shaped association between BMI and POAF^[14, 18]^. Second, some important factors associated with the risk of AF, such as chronic obstructive pulmonary disease (COPD), smoking, diabetes, and left atrial diameter (LAD), were reported to have a higher prevalence in individuals with obesity and those undergoing cardiac surgery. Nevertheless, a univariate meta-analysis cannot account for such confounders in the relationship between obesity and POAF. Third, to date, no comprehensive study has quantitatively assessed the exposure-effect relationship between BMI and POAF. The shape of the exposure–effect curve and whether being overweight increases the risk of POAF are still unclear. Moreover, several studies have also assessed the association between abdominal measures of adiposity, such as waist circumference, and advocated that these measures are a better index for predicting the incidence of POAF^[14, 19]^.

Thus, we conducted a dose-response meta-analysis to quantify the association among body mass index, abdominal adiposity and the incidence of POAF.

## Methods

This study was performed according to Preferred Reporting Items for Systematic reviews and

### Meta-Analyses

Statement (PRISMA) guidelines^[20]^ (**Table S1 in Supplemental Material**). All prospective studies, including RCTs and observational studies (cohort and nested case-control studies), reporting data regarding BMI and POAF were considered eligible for this systematic review. A systematic literature search was conducted using the Cochrane Library, PubMed, and EMBASE databases through December 2019. Two researchers (X.L. and Z-C.T.) independently performed the entire process of this exposure-effect meta-analysis from the literature search and selection to the data analysis. **Table S2 in Supplemental Material** provides a detailed description of the search strategy. All discrepancies were resolved through discussion with each other or consultation with a 3rd reviewer (H-K). We used the method described by Greenland and Longnecker^[21]^ to estimate the study-specific linear trends and 95% CIs from the natural logs of the RRs and CIs across categories of BMI. The robust error meta-regression method was used to fit the nonlinear exposure-effect meta-analysis of BMI and POAF^[22, 23]^. All statistical analyses were performed using Review Manager (RevMan) version 5.3 (The Cochrane Collaboration 2014; Nordic Cochrane Center Copenhagen, Denmark) and Stata software (Version 14.0, Stata Corp LP, College Station, Texas, USA). P<0.05 was considered statistically significant. The full details of the literature search strategy, study selection criteria, quality assessment, and statistical analysis are reported in the **Supplemental Methods in the Supplemental Material.** Additionally, this study was registered with PROSPERO (international prospective register of systematic reviews) under registration number CRD42019128770.

## Results

### Study selection

As shown in **Figure 1**, a total of 655 articles were initially identified, including 189 duplicates. After screening the titles and abstracts, 55 articles remained for the detailed full-text screening. All articles excluded after the full-text review are listed in **Table S3 in the Supplemental Material.** Finally, a total of 30 studies were included. Thirty-two studies were included in the analysis of BMI, and 2 studies were included in the analysis of waist circumference. Seventeen articles reported AF after CABG, two articles reported AF after valve surgery, and eight articles reported the total AF after various cardiac surgeries.

**Figure 1.**
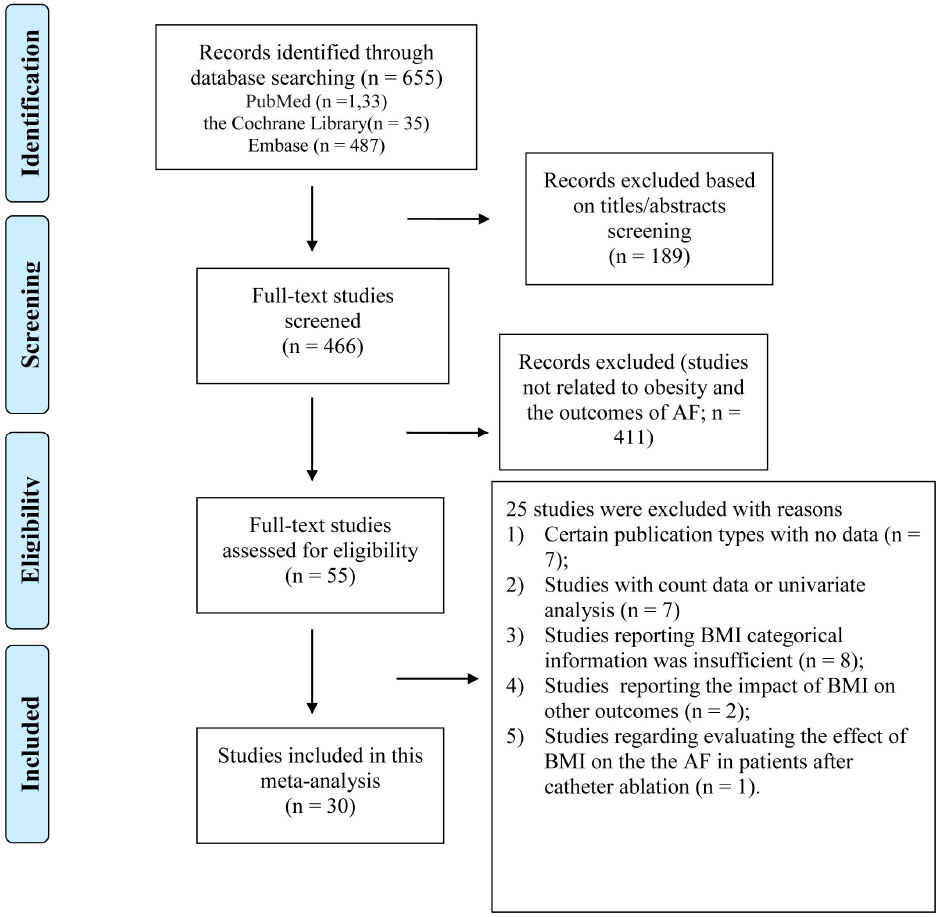
Overview of the research strategy. RR, risk ratio.

### Study characteristics and quality

**Table 1** shows the detailed characteristics of the included studies. Overall, these studies were published between 1996 and 2017. Among the included studies, the sample sizes ranged from 92 to 18,517 with a total of 139,302 individuals. The mean age varied from 56 to 68 years. Fourteen studies^[9, 11, 13, 14, 16, 18, 24-31]^ were from North America (the US and Canada), six studies^[15, 32-36]^ were from Europe, two studies^[10, 12]^ were from Oceania, and two studies^[19, 37]^ were from Asia. Sixteen studies reported the BMI as a categorical variable, eight studies reported the BMI as a continuous variable, and two articles reported the BMI as both a categorical and continuous variable.

**Table 1.**
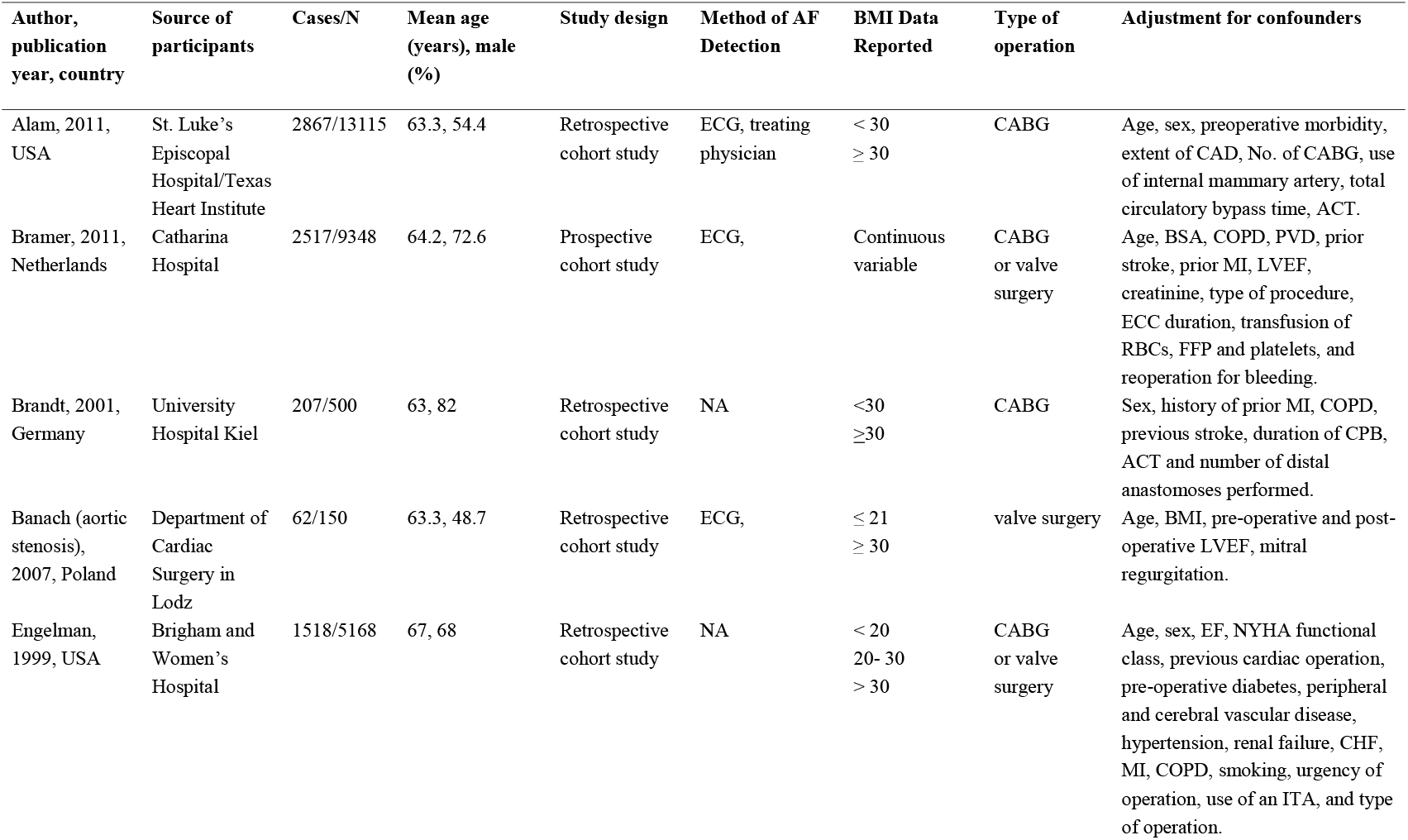
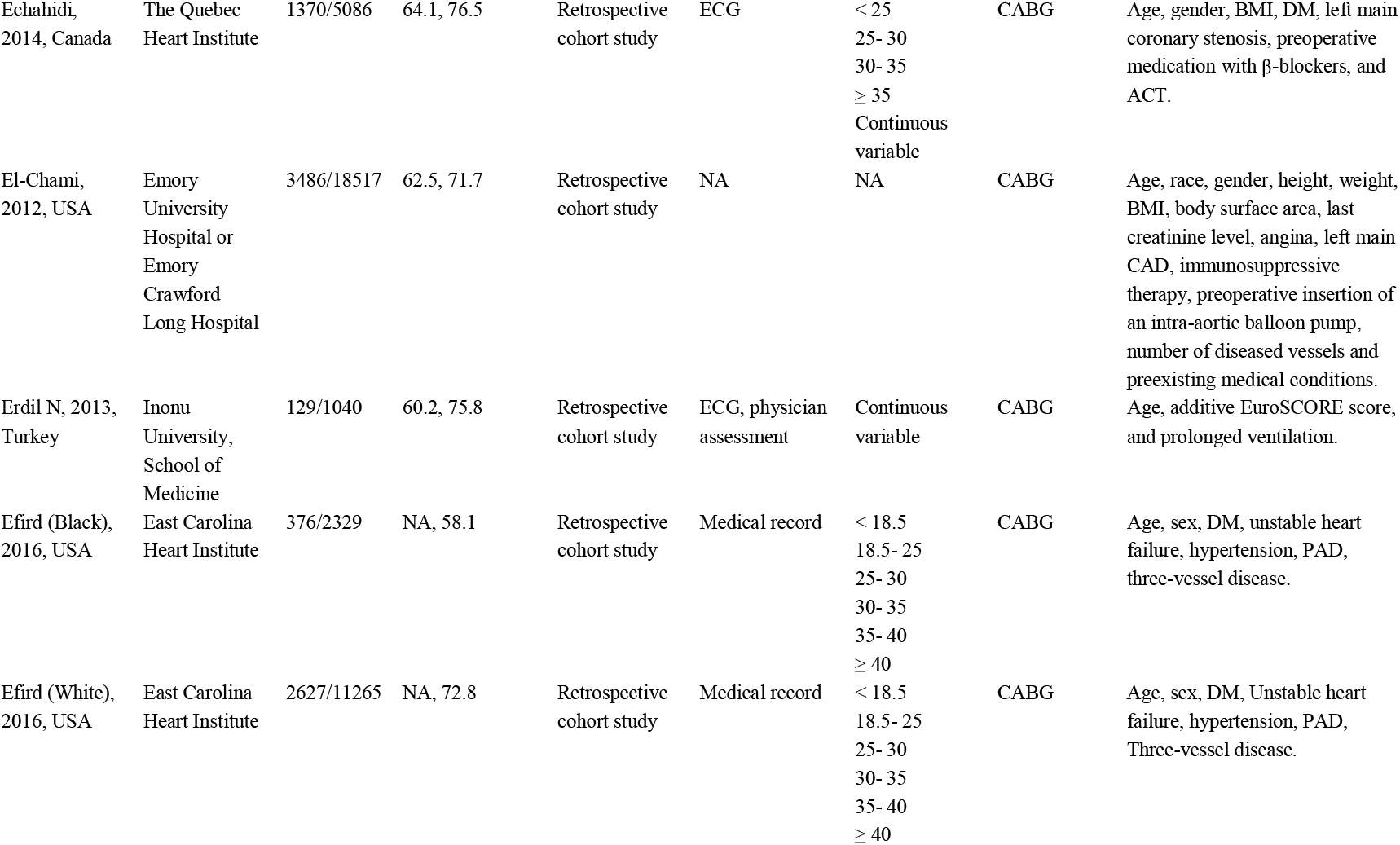
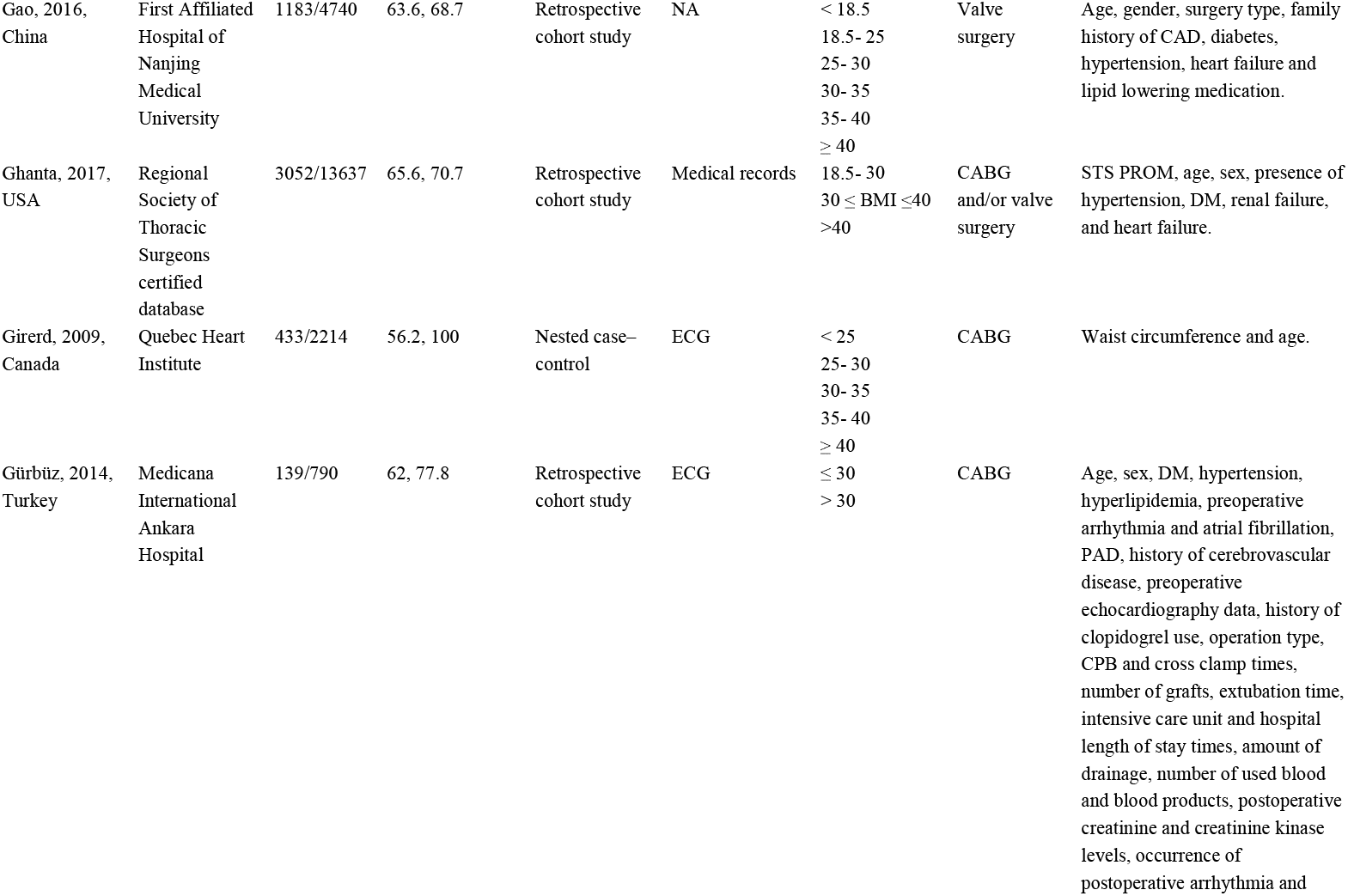
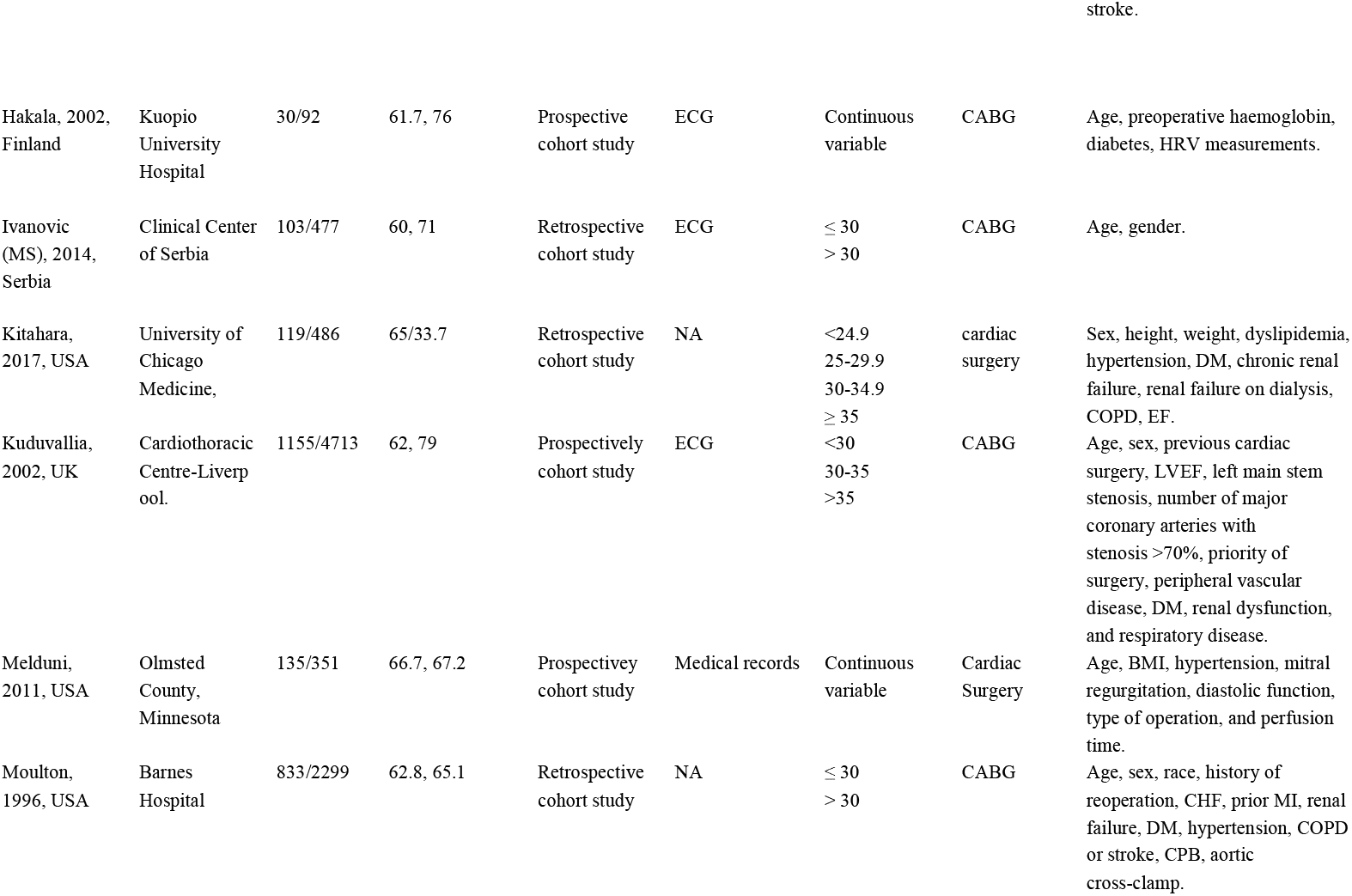
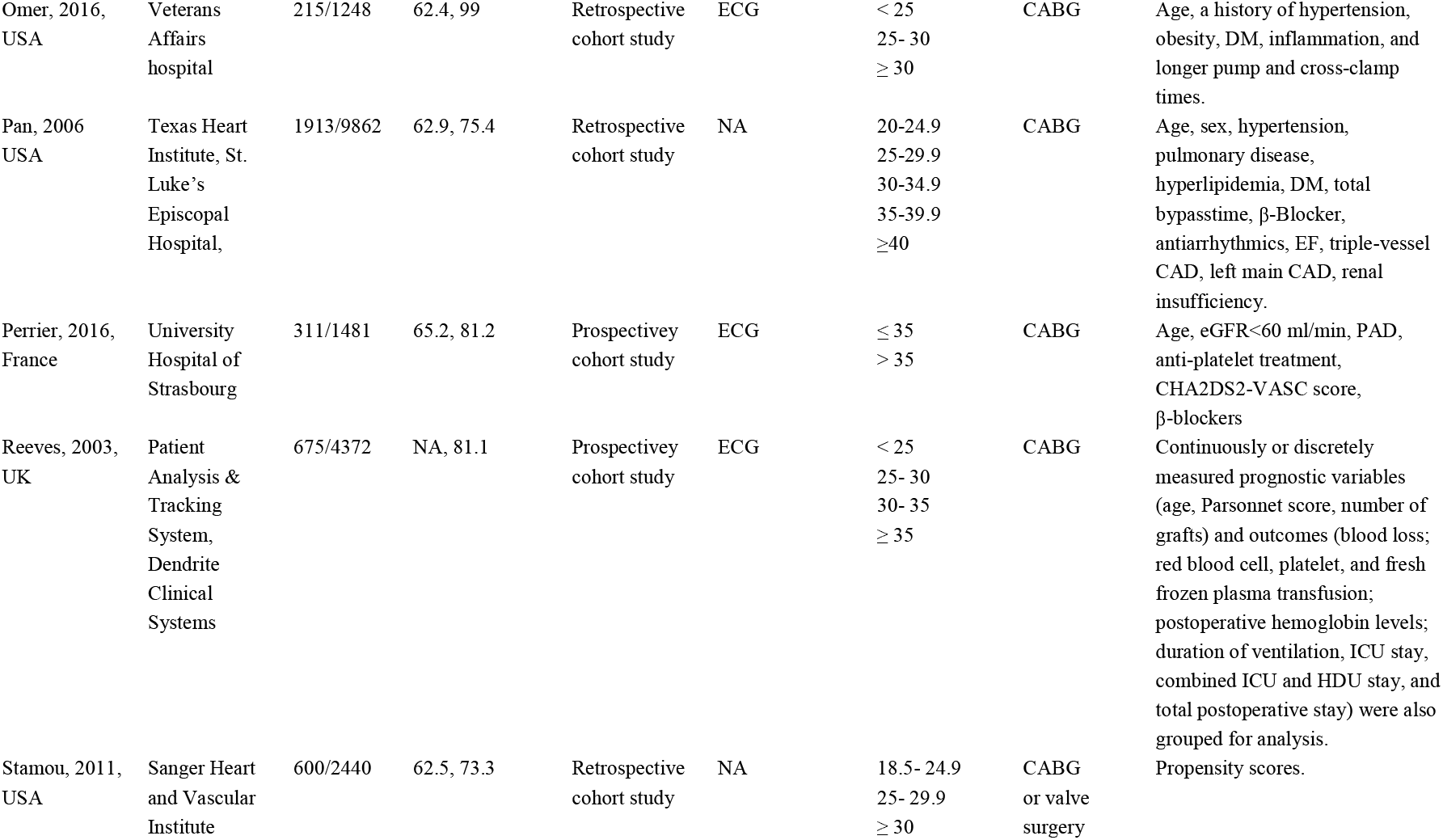
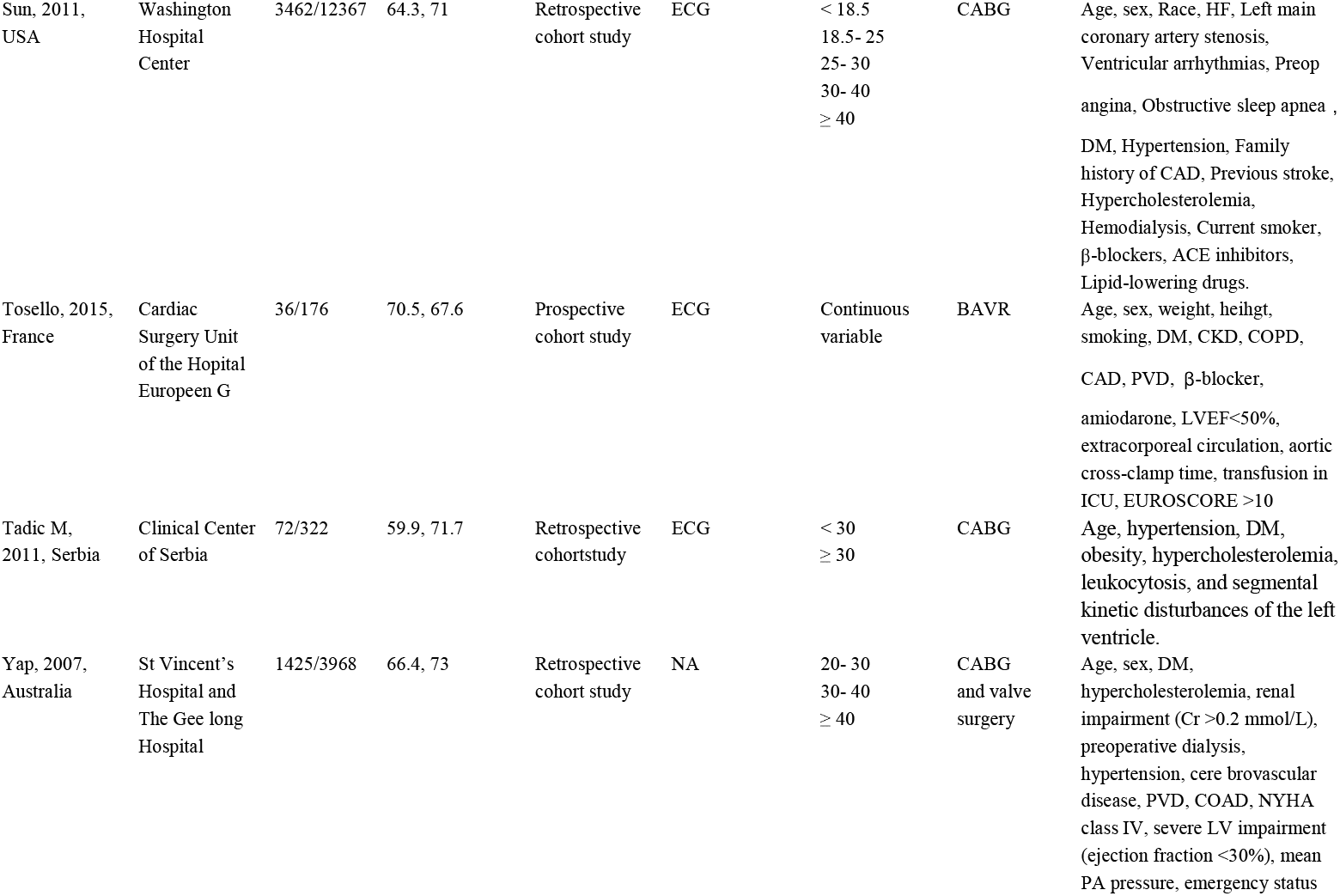
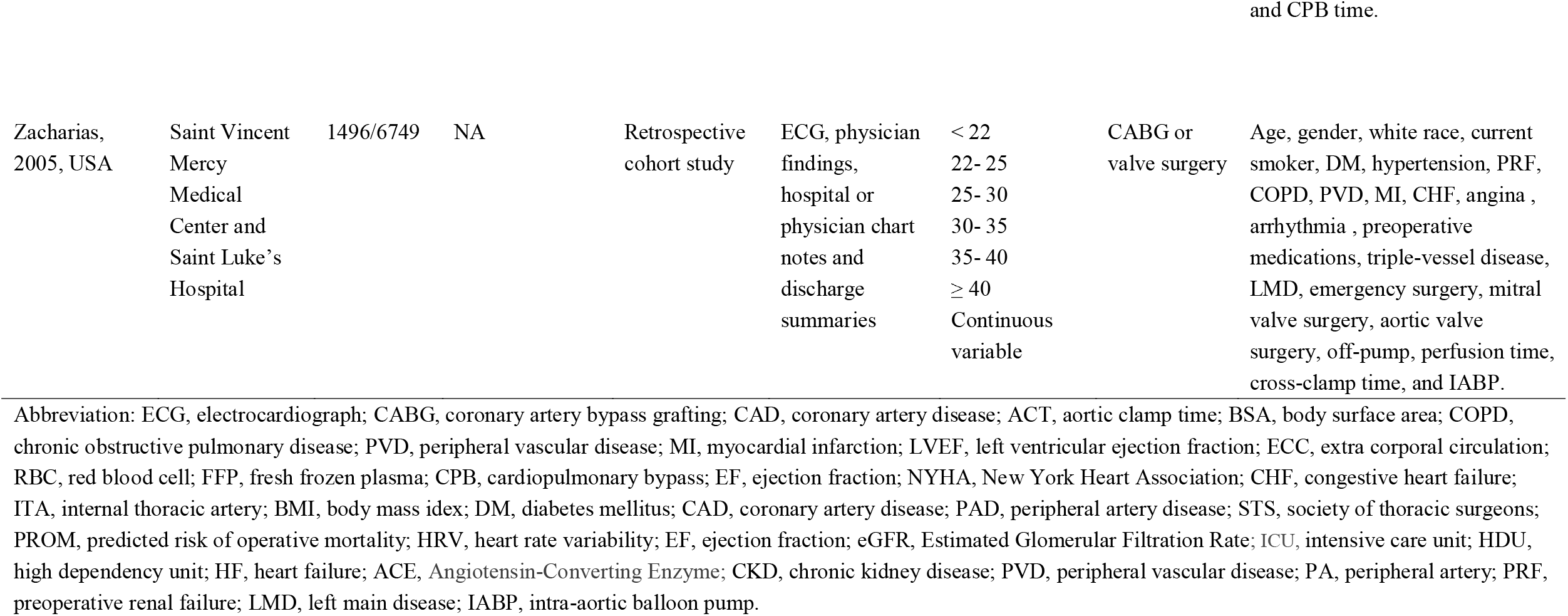
Basic characteristics of the 30 articles included in the meta-analysis.

| **Author, publication year, country** | **Source of participants** | **Cases/N** | **Mean age (years), male (%)** | **Study design** | **Method of AF Detection** | **BMI Data Reported** | **Type of operation** | **Adjustment for confounders** |
| --- | --- | --- | --- | --- | --- | --- | --- | --- |
| Alam, 2011, USA | St. Luke's Episcopal Hospital/Texas Heart Institute | 2867/13115 | 63.3, 54.4 | Retrospective cohort study | ECG, treating physician | < 30<br>≥ 30 | CABG | Age, sex, preoperative morbidity, extent of CAD, No. of CABG, use of internal mammary artery, total circulatory bypass time, ACT. |
| Bramer, 2011, Netherlands | Catharina Hospital | 2517/9348 | 64.2, 72.6 | Prospective cohort study | ECG, | Continuous variable | CABG or valve surgery | Age, BSA, COPD, PVD, prior stroke, prior MI, LVEF, creatinine, type of procedure, ECC duration, transfusion of RBCs, FFP and platelets, and reoperation for bleeding. |
| Brandt, 2001, Germany | University Hospital Kiel | 207/500 | 63, 82 | Retrospective cohort study | NA | <30<br>≥30 | CABG | Sex, history of prior MI, COPD, previous stroke, duration of CPB, ACT and number of distal anastomoses performed. |
| Banach (aortic stenosis), 2007, Poland | Department of Cardiac Surgery in Lodz | 62/150 | 63.3, 48.7 | Retrospective cohort study | ECG, | ≤ 21<br>≥ 30 | valve surgery | Age, BMI, pre-operative and post-operative LVEF, mitral regurgitation. |
| Engelman, 1999, USA | Brigham and Women's Hospital | 1518/5168 | 67, 68 | Retrospective cohort study | NA | < 20<br>20- 30<br>> 30 | CABG or valve surgery | Age, sex, EF, NYHA functional class, previous cardiac operation, pre-operative diabetes, peripheral and cerebral vascular disease, hypertension, renal failure, CHF, MI, COPD, smoking, urgency of operation, use of an ITA, and type of operation. |
| Echahidi, 2014, Canada | The Quebec Heart Institute | 1370/5086 | 64.1, 76.5 | Retrospective cohort study | ECG | < 25<br>25- 30<br>30- 35<br>≥ 35<br>Continuous variable | CABG | Age, gender, BMI, DM, left main coronary stenosis, preoperative medication with β-blockers, and ACT. |
| El-Chami, 2012, USA | Emory University Hospital or Emory Crawford Long Hospital | 3486/18517 | 62.5, 71.7 | Retrospective cohort study | NA | NA | CABG | Age, race, gender, height, weight, BMI, body surface area, last creatinine level, angina, left main CAD, immunosuppressive therapy, preoperative insertion of an intra-aortic balloon pump, number of diseased vessels and preexisting medical conditions. |
| Erdil N, 2013, Turkey | Inonu University, School of Medicine | 129/1040 | 60.2, 75.8 | Retrospective cohort study | ECG, physician assessment | Continuous variable | CABG | Age, additive EuroSCORE score, and prolonged ventilation. |
| Efird (Black), 2016, USA | East Carolina Heart Institute | 376/2329 | NA, 58.1 | Retrospective cohort study | Medical record | < 18.5<br>18.5- 25<br>25- 30<br>30- 35<br>35- 40<br>≥ 40 | CABG | Age, sex, DM, unstable heart failure, hypertension, PAD, three-vessel disease. |
| Efird (White), 2016, USA | East Carolina Heart Institute | 2627/11265 | NA, 72.8 | Retrospective cohort study | Medical record | < 18.5<br>18.5- 25<br>25- 30<br>30- 35<br>35- 40<br>≥ 40 | CABG | Age, sex, DM, Unstable heart failure, hypertension, PAD, Three-vessel disease. |
| Gao, 2016,<br>China | First Affiliated<br>Hospital of<br>Nanjing<br>Medical<br>University | 1183/4740 | 63.6, 68.7 | Retrospective<br>cohort study | NA | < 18.5<br>18.5- 25<br>25- 30<br>30- 35<br>35- 40<br>≥ 40 | Valve<br>surgery | Age, gender, surgery type, family<br>history of CAD, diabetes,<br>hypertension, heart failure and<br>lipid lowering medication. |
| Ghanta, 2017,<br>USA | Regional<br>Society of<br>Thoracic<br>Surgeons<br>certified<br>database | 3052/13637 | 65.6, 70.7 | Retrospective<br>cohort study | Medical records | 18.5- 30<br>30 ≤ BMI ≤ 40<br>>40 | CABG<br>and/or valve<br>surgery | STS PROM, age, sex, presence of<br>hypertension, DM, renal failure,<br>and heart failure. |
| Girerd, 2009,<br>Canada | Quebec Heart<br>Institute | 433/2214 | 56.2, 100 | Nested case–<br>control | ECG | < 25<br>25- 30<br>30- 35<br>35- 40<br>≥ 40 | CABG | Waist circumference and age. |
| Gürbüz, 2014,<br>Turkey | Medicana<br>International<br>Ankara<br>Hospital | 139/790 | 62, 77.8 | Retrospective<br>cohort study | ECG | ≤ 30<br>> 30 | CABG | Age, sex, DM, hypertension,<br>hyperlipidemia, preoperative<br>arrhythmia and atrial fibrillation,<br>PAD, history of cerebrovascular<br>disease, preoperative<br>echocardiography data, history of<br>clopidogrel use, operation type,<br>CPB and cross clamp times,<br>number of grafts, extubation time,<br>intensive care unit and hospital<br>length of stay times, amount of<br>drainage, number of used blood<br>and blood products, postoperative<br>creatinine and creatinine kinase<br>levels, occurrence of<br>postoperative arrhythmia and stroke. |
| Hakala, 2002, Finland | Kuopio University Hospital | 30/92 | 61.7, 76 | Prospective cohort study | ECG | Continuous variable | CABG | Age, preoperative haemoglobin, diabetes, HRV measurements. |
| Ivanovic (MS), 2014, Serbia | Clinical Center of Serbia | 103/477 | 60, 71 | Retrospective cohort study | ECG | ≤ 30<br>> 30 | CABG | Age, gender. |
| Kitahara, 2017, USA | University of Chicago Medicine, | 119/486 | 65/33.7 | Retrospective cohort study | NA | <24.9<br>25-29.9<br>30-34.9<br>≥ 35 | cardiac surgery | Sex, height, weight, dyslipidemia, hypertension, DM, chronic renal failure, renal failure on dialysis, COPD, EF. |
| Kuduvallia, 2002, UK | Cardiothoracic Centre-Liverpool. | 1155/4713 | 62, 79 | Prospectively cohort study | ECG | <30<br>30-35<br>>35 | CABG | Age, sex, previous cardiac surgery, LVEF, left main stem stenosis, number of major coronary arteries with stenosis >70%, priority of surgery, peripheral vascular disease, DM, renal dysfunction, and respiratory disease. |
| Melduni, 2011, USA | Olmsted County, Minnesota | 135/351 | 66.7, 67.2 | Prospective cohort study | Medical records | Continuous variable | Cardiac Surgery | Age, BMI, hypertension, mitral regurgitation, diastolic function, type of operation, and perfusion time. |
| Moulton, 1996, USA | Barnes Hospital | 833/2299 | 62.8, 65.1 | Retrospective cohort study | NA | ≤ 30<br>> 30 | CABG | Age, sex, race, history of reoperation, CHF, prior MI, renal failure, DM, hypertension, COPD or stroke, CPB, aortic cross-clamp. |
| Omer, 2016,<br>USA | Veterans<br>Affairs<br>hospital | 215/1248 | 62.4, 99 | Retrospective<br>cohort study | ECG | < 25<br>25- 30<br>≥ 30 | CABG | Age, a history of hypertension, obesity, DM, inflammation, and longer pump and cross-clamp times. |
| Pan, 2006<br>USA | Texas Heart<br>Institute, St.<br>Luke's<br>Episcopal<br>Hospital, | 1913/9862 | 62.9, 75.4 | Retrospective<br>cohort study | NA | 20-24.9<br>25-29.9<br>30-34.9<br>35-39.9<br>≥40 | CABG | Age, sex, hypertension, pulmonary disease, hyperlipidemia, DM, total bypasstime, β-Blocker, antiarrhythmics, EF, triple-vessel CAD, left main CAD, renal insufficiency. |
| Perrier, 2016,<br>France | University<br>Hospital of<br>Strasbourg | 311/1481 | 65.2, 81.2 | Prospectivey<br>cohort study | ECG | ≤ 35<br>> 35 | CABG | Age, eGFR<60 ml/min, PAD, anti-platelet treatment, CHA2DS2-VASC score, β-blockers |
| Reeves, 2003,<br>UK | Patient<br>Analysis &<br>Tracking<br>System,<br>Dendrite<br>Clinical<br>Systems | 675/4372 | NA, 81.1 | Prospectivey<br>cohort study | ECG | < 25<br>25- 30<br>30- 35<br>≥ 35 | CABG | Continuously or discretely measured prognostic variables (age, Parsonnet score, number of grafts) and outcomes (blood loss; red blood cell, platelet, and fresh frozen plasma transfusion; postoperative hemoglobin levels; duration of ventilation, ICU stay, combined ICU and HDU stay, and total postoperative stay) were also grouped for analysis. |
| Stamou, 2011,<br>USA | Sanger Heart<br>and Vascular<br>Institute | 600/2440 | 62.5, 73.3 | Retrospective<br>cohort study | NA | 18.5- 24.9<br>25- 29.9<br>≥ 30 | CABG<br>or valve<br>surgery | Propensity scores. |
| Sun, 2011, USA | Washington Hospital Center | 3462/12367 | 64.3, 71 | Retrospective cohort study | ECG | < 18.5<br>18.5- 25<br>25- 30<br>30- 40<br>≥ 40 | CABG | Age, sex, Race, HF, Left main coronary artery stenosis, Ventricular arrhythmias, Preop angina, Obstructive sleep apnea , DM, Hypertension, Family history of CAD, Previous stroke, Hypercholesterolemia, Hemodialysis, Current smoker, β-blockers, ACE inhibitors, Lipid-lowering drugs. |
| Tosello, 2015, France | Cardiac Surgery Unit of the Hopital Europeen G | 36/176 | 70.5, 67.6 | Prospective cohort study | ECG | Continuous variable | BAVR | Age, sex, weight, heihgt, smoking, DM, CKD, COPD, CAD, PVD, β-blocker, amiodarone, LVEF<50%, extracorporeal circulation, aortic cross-clamp time, transfusion in ICU, EUROSCORE >10 |
| Tadic M, 2011, Serbia | Clinical Center of Serbia | 72/322 | 59.9, 71.7 | Retrospective cohortstudy | ECG | < 30<br>≥ 30 | CABG | Age, hypertension, DM, obesity, hypercholesterolemia, leukocytosis, and segmental kinetic disturbances of the left ventricle. |
| Yap, 2007, Australia | St Vincent's Hospital and The Gee long Hospital | 1425/3968 | 66.4, 73 | Retrospective cohort study | NA | 20- 30<br>30- 40<br>≥ 40 | CABG and valve surgery | Age, sex, DM, hypercholesterolemia, renal impairment (Cr >0.2 mmol/L), preoperative dialysis, hypertension, cere brovascular disease, PVD, COAD, NYHA class IV, severe LV impairment (ejection fraction <30%), mean PA pressure, emergency status |
| Zacharias,<br>2005, USA | Saint Vincent<br>Mercy<br>Medical<br>Center and<br>Saint Luke's<br>Hospital | 1496/6749 | NA | Retrospective<br>cohort study | ECG, physician<br>findings,<br>hospital or<br>physician chart<br>notes and<br>discharge<br>summaries | < 22<br>22- 25<br>25- 30<br>30- 35<br>35- 40<br>≥ 40<br>Continuous<br>variable | CABG or<br>valve surgery | Age, gender, white race, current<br>smoker, DM, hypertension, PRF,<br>COPD, PVD, MI, CHF, angina ,<br>arrhythmia , preoperative<br>medications, triple-vessel disease,<br>LMD, emergency surgery, mitral<br>valve surgery, aortic valve<br>surgery, off-pump, perfusion time,<br>cross-clamp time, and IABP. |

Abbreviation: ECG, electrocardiograph; CABG, coronary artery bypass grafting; CAD, coronary artery disease; ACT, aortic clamp time; BSA, body surface area; COPD, chronic obstructive pulmonary disease; PVD, peripheral vascular disease; MI, myocardial infarction; LVEF, left ventricular ejection fraction; ECC, extra corporal circulation; RBC, red blood cell; FFP, fresh frozen plasma; CPB, cardiopulmonary bypass; EF, ejection fraction; NYHA, New York Heart Association; CHF, congestive heart failure; ITA, internal thoracic artery; BMI, body mass index; DM, diabetes mellitus; CAD, coronary artery disease; PAD, peripheral artery disease; STS, society of thoracic surgeons; PROM, predicted risk of operative mortality; HRV, heart rate variability; EF, ejection fraction; eGFR, Estimated Glomerular Filtration Rate; ICU, intensive care unit; HDU, high dependency unit; HF, heart failure; ACE, Angiotensin-Converting Enzyme; CKD, chronic kidney disease; PVD, peripheral vascular disease; PA, peripheral artery; PRF, preoperative renal failure; LMD, left main disease; IABP, intra-aortic balloon pump.

The overall reporting quality of the included studies was acceptable. All included studies obtained an NOS≥6 points (**Table S4 in Supplemental Material**).

### Categorical analysis of the effect of BMI on POAF

Fourteen studies grouped the BMI categories according to the World Health Organization criteria; however, only six articles (seven cohorts)^[14, 16, 18, 19, 28, 30]^ reported using normal BMI as the reference group. As shown in **Figure 2**, being underweight was not associated with an increased risk of POAF (RR: 1.44, 95% CI: 0.90–2.30; I^2^*=*29%). In contrast, obesity significantly increased the risk of POAF (RR: 1.39, 95% CI: 1.21–1.61; I^2^=41%). Interestingly, the risk of POAF seemed to gradually increase with the obesity stage (RR of 1.29 for stage I obesity, 1.34 for stage II obesity, and 1.64 for stage III obesity).

**Figure 2.**
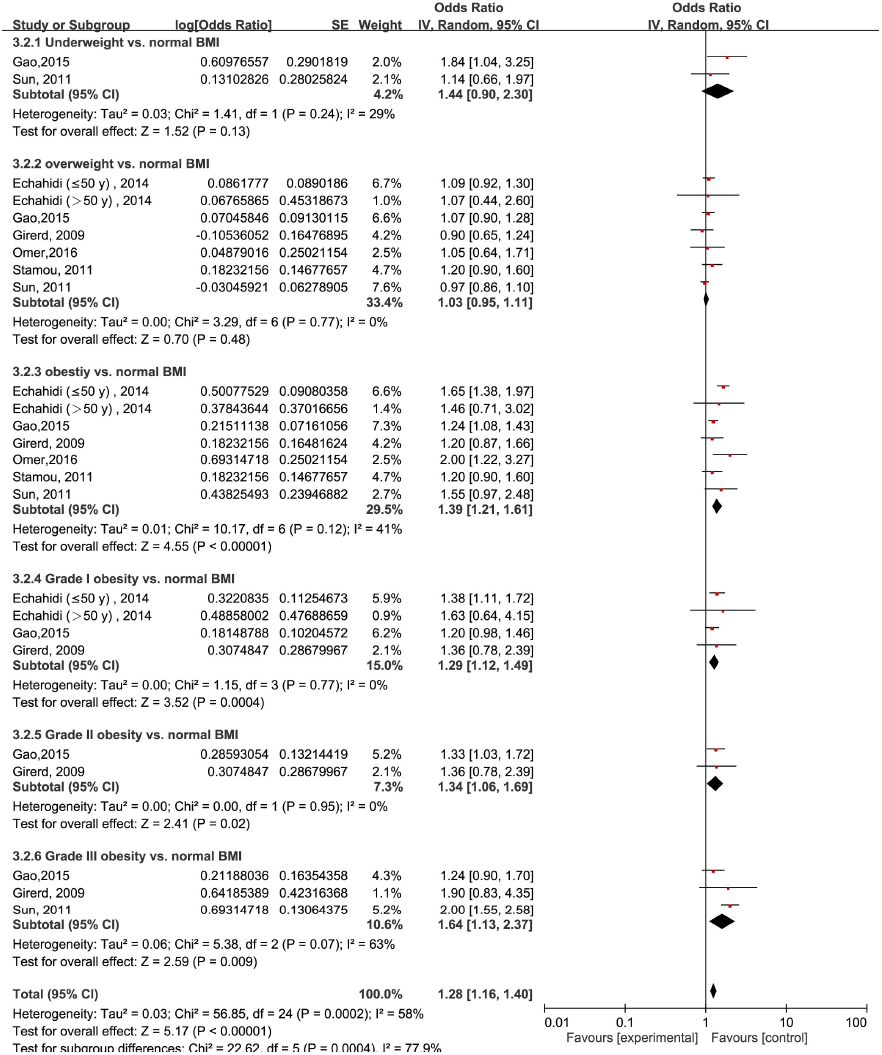
Forest plot of the categorical analysis of the impact of body mass index on POAF. POAF: postoperative atrial fibrillation after cardiac surgery

### Dose-response analysis of the effect of BMI on POAF

Thirty studies^[9-16, 18, 19, 24-30, 32, 33, 35-46]^ involving 32,546 cases/139,302 patients were included in the dose-response analysis of BMI and POAF. The summary RR for a 5-unit increase in BMI was 1.09 (95% CI: 1.05-1.12) with each weight not exceeding 7%. Significant heterogeneity (I^2^=83%) (Figure 3) was found across the studies. In the sensitivity analyses excluding the largest weighted study, the pooled RR ranged from 1.09 (95% CI: 1.05-1.12) to 1.10 (95% CI: 1.06-1.14, I^2^=78%). Additionally, the pooled results were not significantly changed when omitting one study at a time (**Figure S1 in the Supplemental Material**). There was no evidence of nonlinearity (P=0.44) in the relationship between BMI and POAF **(Figure 4)**. The nonlinear curve showed that obesity, but not overweight, significantly increased the risk of POAF compared with the patients with a normal BMI **(Figure 4). Table 3** displays the RR estimates from the nonlinear exposure-effect analysis of selected BMI values; these values were derived from the nonlinear figures.

**Table 2.**
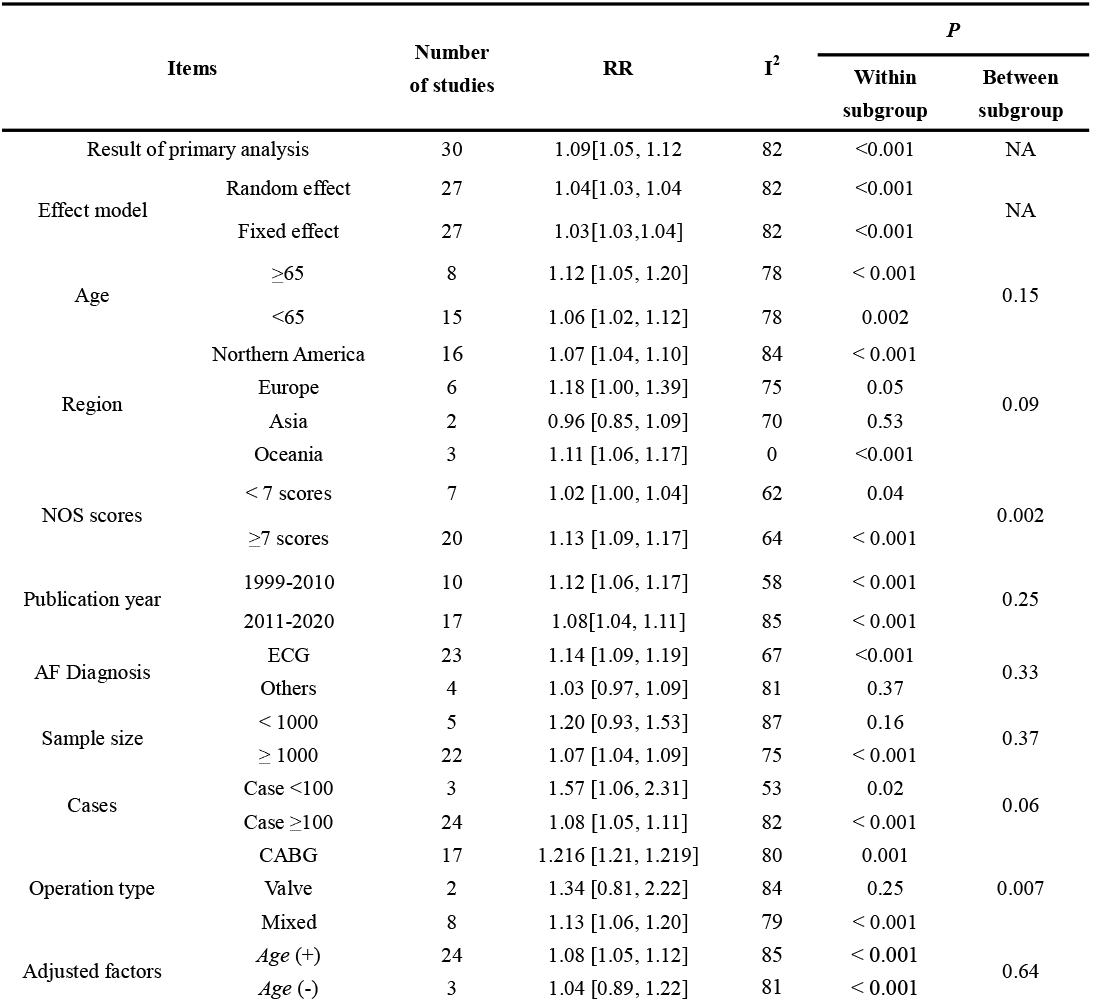
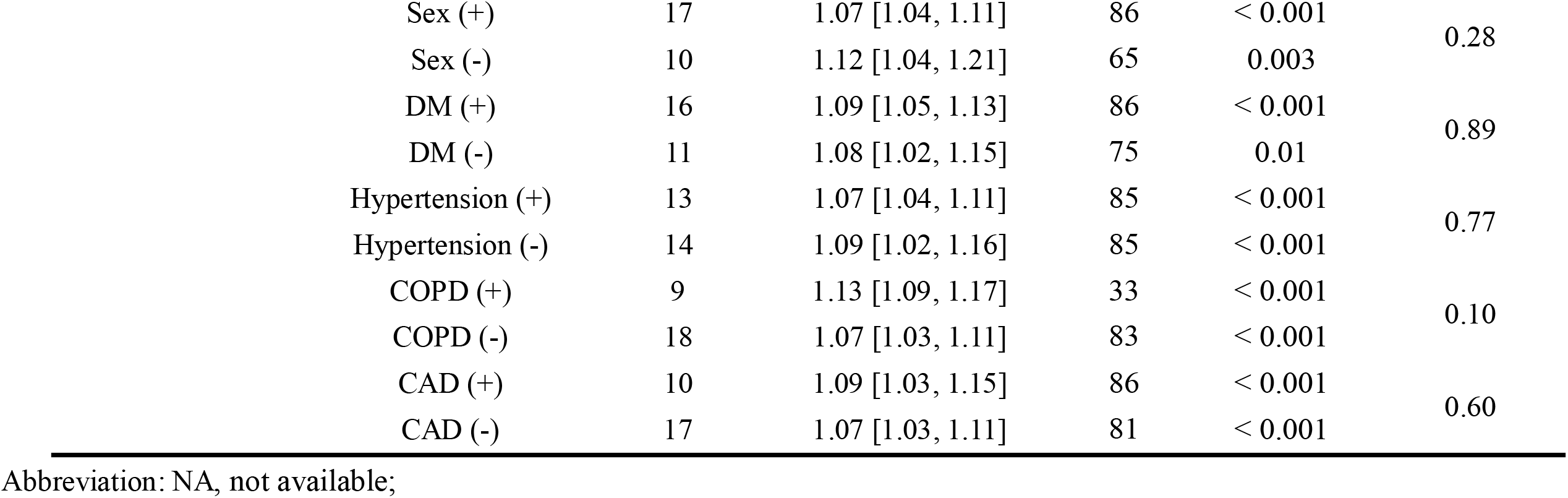
Subgroup analysis of body mass index and post-cardiac operation atrial fibrillation.

| Items |  | Number<br>of studies | RR | I<sup>2</sup> | *P* |  |
| --- | --- | --- | --- | --- | --- | --- |
|  |  |  |  |  | Within<br>subgroup | Between<br>subgroup |
| Result of primary analysis |  | 30 | 1.09[1.05, 1.12] | 82 | <0.001 | NA |
| Effect model | Random effect | 27 | 1.04[1.03, 1.04] | 82 | <0.001 | NA |
|  | Fixed effect | 27 | 1.03[1.03,1.04] | 82 | <0.001 |  |
| Age | ≥65 | 8 | 1.12 [1.05, 1.20] | 78 | < 0.001 | 0.15 |
|  | <65 | 15 | 1.06 [1.02, 1.12] | 78 | 0.002 |  |
| Region | Northern America | 16 | 1.07 [1.04, 1.10] | 84 | < 0.001 | 0.09 |
|  | Europe | 6 | 1.18 [1.00, 1.39] | 75 | 0.05 |  |
|  | Asia | 2 | 0.96 [0.85, 1.09] | 70 | 0.53 |  |
|  | Oceania | 3 | 1.11 [1.06, 1.17] | 0 | <0.001 |  |
| NOS scores | < 7 scores | 7 | 1.02 [1.00, 1.04] | 62 | 0.04 | 0.002 |
|  | ≥7 scores | 20 | 1.13 [1.09, 1.17] | 64 | < 0.001 |  |
| Publication year | 1999-2010 | 10 | 1.12 [1.06, 1.17] | 58 | < 0.001 | 0.25 |
|  | 2011-2020 | 17 | 1.08[1.04, 1.11] | 85 | < 0.001 |  |
| AF Diagnosis | ECG | 23 | 1.14 [1.09, 1.19] | 67 | <0.001 | 0.33 |
|  | Others | 4 | 1.03 [0.97, 1.09] | 81 | 0.37 |  |
| Sample size | < 1000 | 5 | 1.20 [0.93, 1.53] | 87 | 0.16 | 0.37 |
|  | ≥ 1000 | 22 | 1.07 [1.04, 1.09] | 75 | < 0.001 |  |
| Cases | Case <100 | 3 | 1.57 [1.06, 2.31] | 53 | 0.02 | 0.06 |
|  | Case ≥100 | 24 | 1.08 [1.05, 1.11] | 82 | < 0.001 |  |
| Operation type | CABG | 17 | 1.216 [1.21, 1.219] | 80 | 0.001 | 0.007 |
|  | Valve | 2 | 1.34 [0.81, 2.22] | 84 | 0.25 |  |
|  | Mixed | 8 | 1.13 [1.06, 1.20] | 79 | < 0.001 |  |
| Adjusted factors | Age (+) | 24 | 1.08 [1.05, 1.12] | 85 | < 0.001 | 0.64 |
|  | Age (-) | 3 | 1.04 [0.89, 1.22] | 81 | < 0.001 |  |

|  |  |  |  |  |  |
| --- | --- | --- | --- | --- | --- |
| Sex (+) | 17 | 1.07 [1.04, 1.11] | 86 | < 0.001 | 0.28 |
| Sex (-) | 10 | 1.12 [1.04, 1.21] | 65 | 0.003 |  |
| DM (+) | 16 | 1.09 [1.05, 1.13] | 86 | < 0.001 | 0.89 |
| DM (-) | 11 | 1.08 [1.02, 1.15] | 75 | 0.01 |  |
| Hypertension (+) | 13 | 1.07 [1.04, 1.11] | 85 | < 0.001 | 0.77 |
| Hypertension (-) | 14 | 1.09 [1.02, 1.16] | 85 | < 0.001 |  |
| COPD (+) | 9 | 1.13 [1.09, 1.17] | 33 | < 0.001 | 0.10 |
| COPD (-) | 18 | 1.07 [1.03, 1.11] | 83 | < 0.001 |  |
| CAD (+) | 10 | 1.09 [1.03, 1.15] | 86 | < 0.001 | 0.60 |
| CAD (-) | 17 | 1.07 [1.03, 1.11] | 81 | < 0.001 |  |

Abbreviation: NA, not available;

**Table 3:**
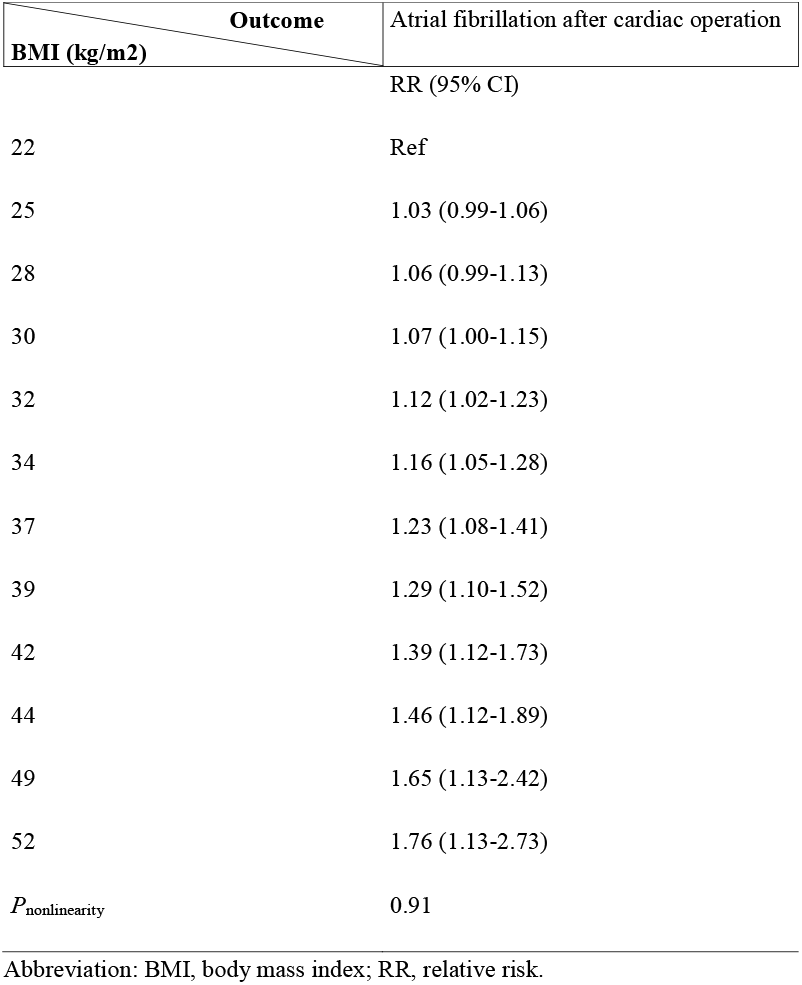
Relative risks between BMI and atrial fibrillation incident after cardiac operation, from the nonlinear dose-response analysis

| BMI (kg/m<sup>2</sup>) | Outcome |
| --- | --- |
|  | Atrial fibrillation after cardiac operation |
|  | RR (95% CI) |
| 22 | Ref |
| 25 | 1.03 (0.99-1.06) |
| 28 | 1.06 (0.99-1.13) |
| 30 | 1.07 (1.00-1.15) |
| 32 | 1.12 (1.02-1.23) |
| 34 | 1.16 (1.05-1.28) |
| 37 | 1.23 (1.08-1.41) |
| 39 | 1.29 (1.10-1.52) |
| 42 | 1.39 (1.12-1.73) |
| 44 | 1.46 (1.12-1.89) |
| 49 | 1.65 (1.13-2.42) |
| 52 | 1.76 (1.13-2.73) |
| $P_{\text{nonlinearity}}$ | 0.91 |

Abbreviation: BMI, body mass index; RR, relative risk.

**Figure 3.**
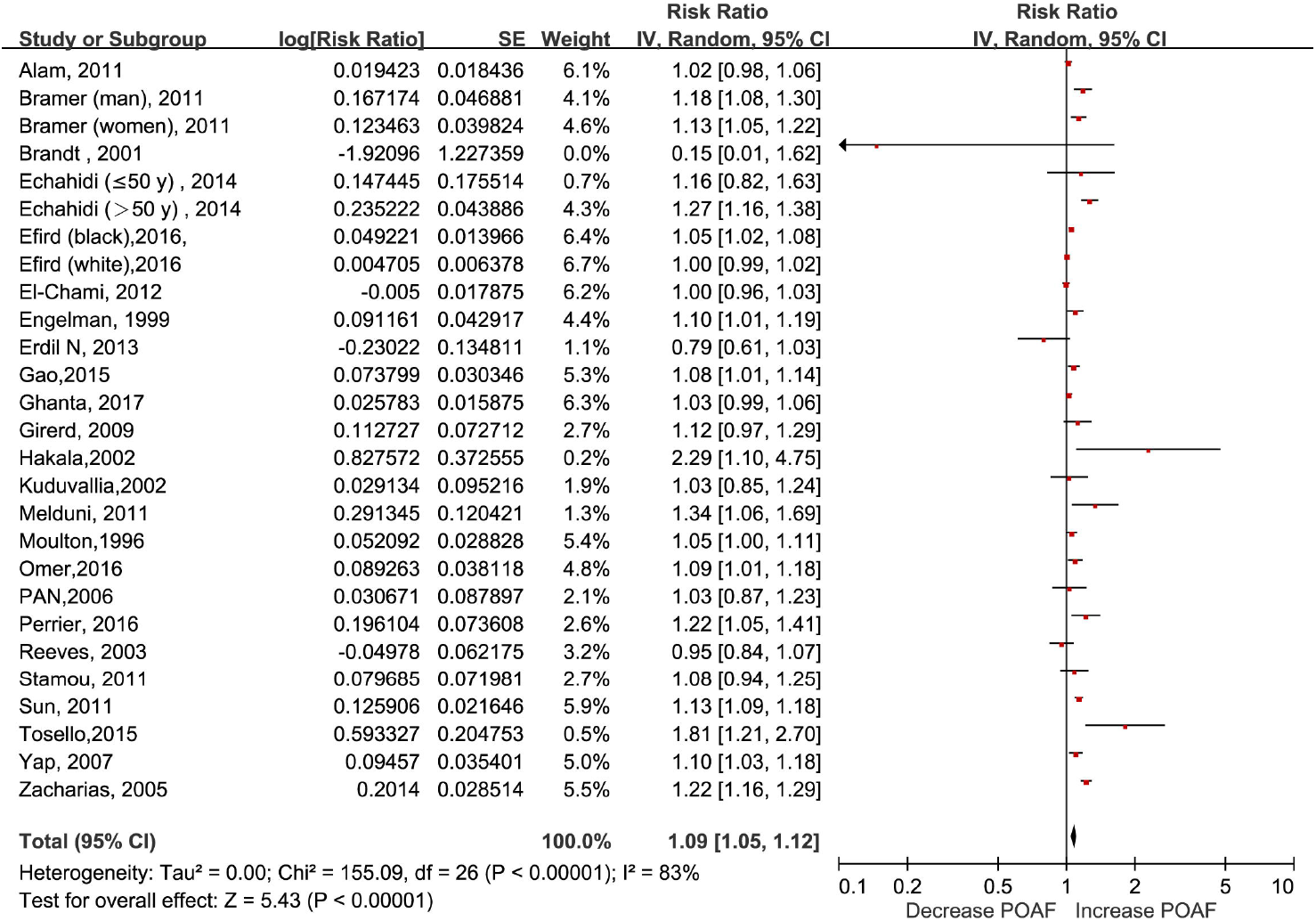
Forest plot of the association between body mass index and POAF and exposure-effect analysis, per 5 units. POAF: postoperative atrial fibrillation after cardiac surgery

**Figure 4.**
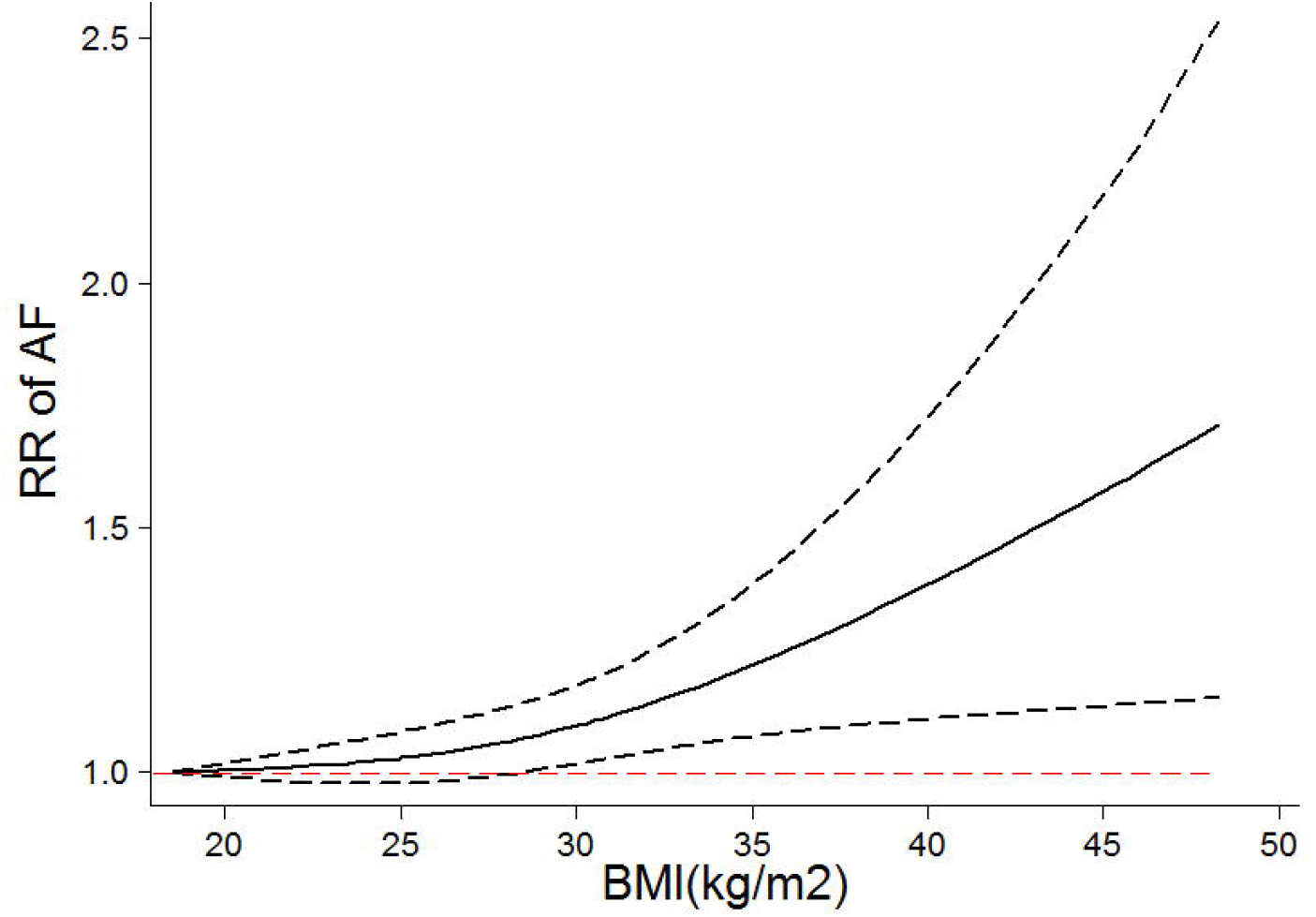
Nonlinear exposure-effect analysis of body mass index and POAF. The solid and dashed lines represent the estimated relative risk and the 95% confidence interval, respectively. POAF: postoperative atrial fibrillation after cardiac surgery

We further performed a subgroup analysis by type of cardiac operation. Seventeen articles^[13-16, 18, 24, 25, 28, 29, 34, 35, 39, 42]^ reported the association in coronary artery bypass graft (CABG), two articles^[19, 36]^ reported the association in valve surgery, and seven articles^[9-13, 30, 31]^ reported the association in combined types of cardiac surgery. The summary RRs for a 5-unit increment in BMI in the CABG group, valve surgery group, and combined cardiac surgery group were 1.216 (95% CI: 1.21-1.219, I^2^=80%), 1.34 (95% CI: 0.81-2.22, I^2^=84%), and 1.13 (95% CI: 1.06-1.20, I^2^=79%), respectively **(Table 2).** Moreover, there was some evidence of a J-shaped relationship between POAF in the CABG group (P_nonlinearity_=0.0027), with the risk of POAF significantly increasing at a BMI of 30 and rising steeply at higher BMI levels. However, the curve was much flatter in the combined cardiac surgery group **(Figure 5A-B)**.

**Figure 5.**
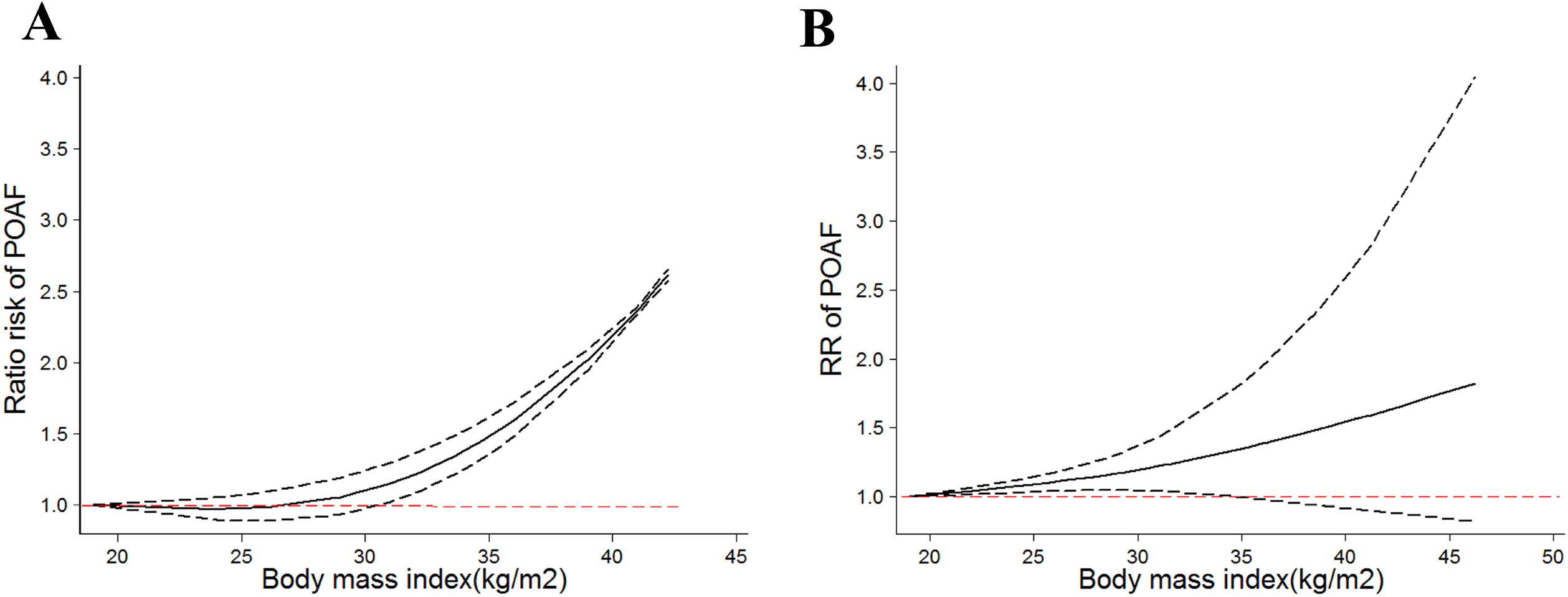
Body mass index and POAF stratified by types of cardiac surgery and nonlinear exposure-effect analysis. The solid and dashed lines represent the estimated relative risk and the 95% confidence interval, respectively. A: coronary artery bypass graft; B: combined cardiac surgery. POAF: postoperative atrial fibrillation after cardiac surgery

### Waist circumference obesity and POAF

Two prospective studies^[14, 39]^ reported an analysis of the association between waist circumference and the risk of AF after CABG and included 536 cases among 2,691 participants. Abdominal obesity was associated with an increased risk of POAF (RR: 1.55, 95% CI: 1.26-1.90) with no evidence of heterogeneity (I^2^=0%) **(Figure 6).**

**Figure 6.**
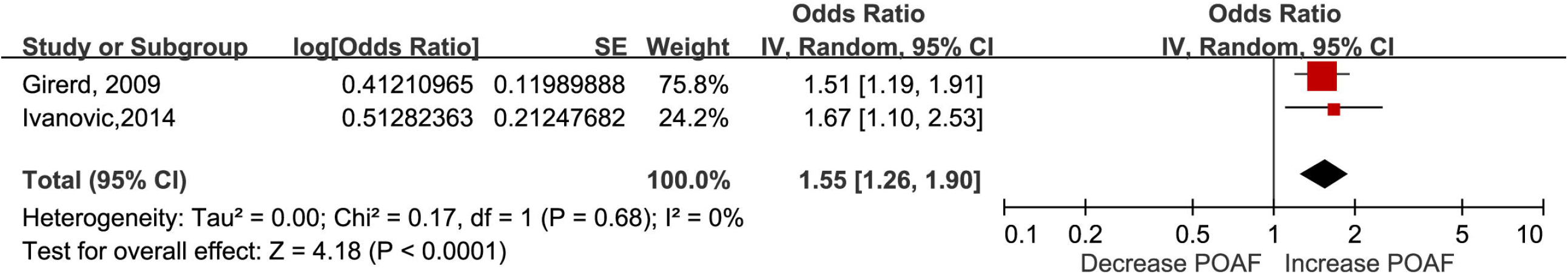
Forest plot of the impact of waist obesity on POAF. POAF: postoperative atrial fibrillation after cardiac surgery

### Subgroup and meta-regression analyses

We conducted a subgroup analysis and a meta-regression by patient characteristics, such as age, region, confounding factors and potential intermediate factors. We found some indication of a stronger relationship between BMI and POAF among the studies with higher NOS scores (**Table 2**). As shown in **Table 2**, the positive association between BMI and the risk of POAF persisted in almost all subgroup analyses by age, region, sample size, study quality and adjustment for clinical confounding factors (e.g., diabetes, hypertension, and age), and there was no evidence of heterogeneity among any of these subgroups in the meta-regression analyses.

### Publication bias

A possible lack of publication bias was indicated by Egger’s and Begg’s tests and the funnel plot (**Figure S2-4 in the Supplemental Material**).

## Discussion

This study presents the first dose-response analysis of the relationship between adiposity and the risk of POAF. By combining 30 prospective studies involving 32,546 cases among 139,302 patients, we found a 9% increased risk of POAF per a 5-unit increase in BMI. Both the categorical analysis and exposure-effect model showed that obesity, but not being overweight or underweight, significantly increased the risk of POAF. Consistently, in the subgroup analysis of CABG, a somewhat J-shaped association was found as the risk significantly increased at a BMI of 30 and steeply rose at higher BMI levels. Finally, to the best of our knowledge, we are also the first to confirm the association between abdominal adiposity and the risk of AF in patients after cardiac surgery. These findings provide a compressive overview of the association between adiposity and POAF.

Although the relationship between obesity and the risk of POAF is not new, the exposure-effect relationship between BMI and POAF is still unclear, and the associations between overweight and the risk of POAF have not been comprehensively assessed. Several prospective cohort studies^[47, 48]^ found that being overweight significantly increased the risk of new onset AF; however, an independent positive relationship in POAF was not confirmed in several large prospective long-term follow-up cohorts after adjusting for clinical confounding variables^[49, 50]^ (e.g., the Framingham Heart Study). Consistently, our categorical analysis and exposure-effect analysis uniformly showed that being overweight did not statically increase the risk of POAF. This result was unsurprising. First, as described by a previously published meta-analysis^[51, 52]^, the magnitude of the association between overall obesity and POAF is minimal, with an RR of 1.12-1.21. Consistently, our results also showed that the summary RR per 5 units of BMI was 9%, further suggesting that the real effect may even be very small in magnitude. Furthermore, the risk factors for POAF are complicated and mainly include transient perioperative factors and a pre-existing condition^[53]^. A large cohort study^[54]^ found that the repeat recurrence of AF in cardiac surgery was higher than that in noncardiac operations, supporting that the role of transient factors (e.g., cross-clamp time and intra-aortic balloon pump) is more significant than that of a pre-existing condition (e.g., obesity or hypertension). Thus, we speculated that the pathological condition caused by overweight might be compensable and insufficient to trigger POAF independently.

The association between obesity and atrial fibrillation is not new in the field of cardiac surgery. However, due to the limitations of univariate analyses and lack of evaluation of other clinical factors or potential intermediates in previously published meta-analyses^[51, 52]^, the multitude of other factors and potential intermediates (e.g., age, smoking, obstructive sleep apnea (OSA), COPD, and hypertension) that may also affect the association between obesity and POAF incidence are still unclear. For example, the association of hypertension with POAF was slightly stronger than that of obesity and was more common in cardiac surgery patients with obesity. Stamou et al ^[30]^ found that 87% of patients with obesity had preoperative hypertension compared to 75% of patients without obesity in a cohort of 2,465 patients undergoing cardiac surgery. Another important intermediate factor is lung disease. For example, a study reported that the incidence of POAF increases with surgical invasiveness from an RR of 2.26 after mediastinal surgery to 8.90 in patients undergoing pneumonectomy, suggesting that a strong association exists between lung disease and POAF^[55]^. More consistently, several studies also showed that COPD was strongly linked to the incidence and progression of AF^[56, 57]^. Another study also showed that POAF was associated primarily with metabolic syndrome (OR, 2.36; p=0.02) rather than BMI in a younger population^[16]^. Our results showed that the positive association persisted in almost all subgroup analyses by age, region, number of cases, and study quality and after adjustment for these abovementioned factors, indicating an independent association between obesity and the risk of POAF.

Notably, not all important clinical confounding factors or potential intermediates were assessed in the present study. For example, OSA was common in patients with obesity and sharply increased with BMI. The link between OSA and new onset incident AF has been confirmed^[58]^. Various studies have found that OSA is independently associated with an increased risk of AF following CABG^[59, 60]^. Thus, some authors have suggested that the positive correlation between obesity and POAF can be explained by OSA. Similarly, LAD enlargement, which is another pathological condition that often coexisted with obesity, was also identified as a crucial factor that independently contributes to the incidence of AF^[61]^. However, few included studies adjusted for OSA or enlarged LAD, and even fewer studies examined the direct link among OSA, LAD and increased POAF. Therefore, the role of OSA and LAD in the relationship between BMI and POAF is still unclear and needs to be further studied.

Notably, BMI is commonly used because it is simple to apply and inexpensive. However, the utility of BMI in evaluating obesity has been criticized because it is indistinguishable from fat. In our results, body fat measured by waist circumference increased the risk of POAF by 55%. This value was much greater than that previously reported in the association between obesity and POAF and the value observed in our study. Although the included study was limited, some authors have highlighted the importance of using multiple measures, such as visceral obesity, in the risk assessment of POAF^[14]^.

Previous studies only analyzed the association between obesity and the POAF incidence in total cardiac surgery^[51, 52]^. The potential difference in the impact of obesity on cardiac surgery has not been examined. In the present study, a significant difference was found in the cardiac surgery subgroup analysis (P of subgroup difference=0.007). Furthermore, a nonlinear expose-effect relationship between BMI and POAF in CABG was observed, with a sharper and steeper curve. In contrast, the exposure-effect curve of the combined types of cardiac surgery subgroup (CABG and valve surgery) was much flatter. This result suggests that the curve of POAF after valve surgery was much lower than that in the CABG group. This result was further reinforced by the linear analysis, which showed a nonsignificant RR (P=0.25) per a 5-unit increase in BMI. However, we should interpret these results with caution. The number of studies reporting obesity and POAF in valve surgery was limited, and the pooled RR had substantial heterogeneity. Moreover, in another cohort^[13]^ consisting of 16% valve surgery and 85% CABG, similar RRs were found across different obesity groups in the analysis between total surgery and isolated CABG only. Therefore, the impact of obesity on CABG and valve surgery is still unclear and should be further studied.

The effect of POAF on secondary outcomes was not assessed in the present study. Previous studies have observed an “obesity paradox” in the outcomes of patients undergoing cardiac surgery, with individuals affected by overweight and class I and II obesity having lower mortality^[30, 62]^. However, recent studies using dose-response methods and more compressive meta-analyses did not find any protective effect or higher mortality among patients with extreme obesity^[63]^. Consistently, another meta-analysis found higher major morbidity and total hospital costs in patients with obesity undergoing cardiac surgery^[31]^. Our results further support this observation of a lack of an obesity paradox; we found that POAF increased with increasing BMI, and morbidly increased POAF by 40% and 120% in the total cardiac surgery and CABG subgroups, respectively. POAF independently predicted stroke^[64]^ and long-term mortality. Thus, regarding obesity, especially morbid obesity, the risk of POAF should be carefully evaluated before cardiac surgery, and specific interventions for the prevention of POAF should be considered. To date, purposeful weight loss has been shown to reverse many changes in cardiac performance and morphology associated with obesity and the incidence and burden of AF^[65, 66]^. However, weight loss might not be as prevalent in the cardiac surgery subgroup because most CABG procedures are emergent and unexpected or conducted in patients with poor cardiac function (patients receiving valve replace). Alternatively, the administration of certain medications was the most commonly used therapy for POAF prophylaxis. Among these medications, β-blockers, amiodarone, and sotalol were proven to be effective against POAF after cardiac surgery. β-blockers were uniformly recommended by the guidelines (class I recommendation)^[67-69]^. According to a Cochrane review meta-analysis, β-blockers significantly reduce the incidence of POAF by 67%^[70]^, and the initiation or continuation of β-blockers is strongly recommended. Amiodarone is recommended for patients with contraindications to β-blockers or (alone or in addition to β-blocker therapy) those who are at a high risk of POAF. Sotalol is recommended for general POAF prophylaxis (AHA/ACC/HRS class IIb recommendation)^[53]^. An RCT found that sotalol was equally effective for prophylaxis of POAF as amiodarone^[71]^. Moreover, sotalol was even more effective than standard β-blocker therapy in patients undergoing cardiac surgery^[71]^. Notably, obesity often coexists with diabetes, an older age, congestive heart failure, and other risk factors, and the risk of POAF might increase by multiple times as a result. Thus, a combination of β-blockers and amiodarone can be considered for POAF prophylaxis in these high-risk patients.

Finally, our findings have important clinical implications for the prevention of POAF as a previous meta-analysis analyzed BMI but did not include other measures of fat in relation to the risk of POAF, and prior studies did not assess the dose-response relationship between BMI and POAF in as great detail as the present analysis. In addition, our results show that obesity was associated with an increased risk of POAF in studies from Europe, North America, and Oceania, suggesting that the prevention of obesity is essential across populations. The current analysis suggests that both general and abdominal adiposity (waist circumference) measures are related to an increased risk of POAF and that being relatively slim, as assessed by BMI and other adiposity measures, may confer the lowest risk of POAF.

### Strengths and limitations

Our meta-analysis has several strengths. First, all included studies were designed as prospective studies, avoiding recall bias and reducing the possibility of selection bias. Second, this meta-analysis included a large number of cohort studies (30 studies, N=139,302) with BMI reported as either a categorical or continuous variable, providing strong statistical power to detect moderate associations. The detailed exposure–effect analyses clarified the shape of the exposure–effect relationship. Third, the positive association between BMI and POAF persisted in different subgroups (e.g., age or region) and after adjusting for confounding factors, confirming the robustness of our findings.

Our study inevitably has several limitations. First, this meta-analysis included observational studies, and bias was not entirely avoided. Measured and unmeasured confounding variables might have influenced our results. However, this limitation cannot be mitigated by the large number of studies and adjustments for coexistent confounding factors in all included studies. Second, significant heterogeneity (I^2^=83%) was observed across the included studies, which might have been derived from between-study differences, such as differences in study design, basic patient characteristics, and analytical strategies. Third, we did not analyze the long-term POAF incidence since most studies only reported the incidence of POAF during the length of hospitalization. However, most POAF has a peak incidence between days 2 and 4 after surgery^[53]^, especially during the first postoperative week. Fourth, we did not assess the effect of BMI on secondary outcomes in patients undergoing a cardiac operation, such as stroke and death, which have been thoroughly studied in previous studies^[31, 63]^.

In conclusion, based on the current evidence, our findings are the first to show that obesity and abdominal adiposity are independently associated with an increased risk of POAF. Being underweight or overweight might not significantly increase the POAF risk. The magnitude of the effect of obesity on AF in patients undergoing valve surgery might be small and needs to be further confirmed.

## Data Availability

All data generated or analyzed during this study are included in this article.

## Sources of funding

This work was supported in part by the National Natural Science Foundation of China (No. 81760050, 81760048) and the Jiangxi Provincial Natural Science Foundation for Youth Scientific Research (No. 20192ACBL21037).

## Conflicts of interest

All authors declare that they have no conflicts of interest.

## Author contributions

X.L. was responsible for the entire project and revised the draft. X.L. and M-L.L. performed the systematic literature review and drafted the first version of the manuscript. All authors participated in the interpretation of the results and prepared the final version of the manuscript.

## Notes

### Competing Interest Statement

The authors have declared no competing interest.

