## supplemental materials for "Dose-response relationship among body mass index, abdominal adiposity and atrial fibrillation in patients undergoing cardiac surgery: a meta-analysis of 30 prospective studies"

**SUPPLEMENTAL METHODS**

**Literature Search**

We systematically searched the Cochrane Library, PubMed, and Embase databases for eligible studies from inception through May 1, 2019. Three groups of keywords or MeSH terms (linked to body mass index, waist circumference, and waist-to-hip ratio, atrial fibrillation and cardiac surgery) were combined using the Boolean operator "and". In addition, we searched the reference lists of other relevant publications to identify further studies. No language restrictions were applied in the whole literature search.

**Study Selection**

Studies were considered eligible if they: (1) designed as prospective studies including post-hoc analysis of randomized controlled trials (RCTs) or observational studies(cohort or nest case-control); (2) reported the adjusted RR for association btewwen body mass index (BMI), waist circumference, and waist-to-hip ratio on atrial fibrillation after cardiac surgery(POAF); (3) made available a quantitative measure of adiposity and the number of POAF cases in each adiposity category for the dose-response analysis. For multiple publications/reports created from the same data, the studies with the l the largest number of POAF cases were included. In addition, certain publication types (e.g., reviews, editorials, letters, conference abstracts, and animal studies), or studies with insufficient data were excluded from this analysis.

**Data Extraction and Quality Assessment**

For each study, the basic characteristics were extracted, mainly including the first author, publication year, geographical location, study type, participants (sex, age, and sample size), duration of follow-up, adjustments for confounders, categories of adiposity and adjusted risk ratios (RRs) with its 95% confidence intervals (CIs) for each adiposity category. If both unadjusted and adjusted RRs existed in one study, we extracted the most completely adjusted one.

Post-hoc analyses of RCTs can be equivalent to observational studies.^1^ Therefore, we used the Newcastle-Ottawa quality assessment scale (NOS) to evaluate the quality for all included studies.^2^ The validated NOS items with a total of 9 stars involved three aspects including the selection of cohorts, the comparability of cohorts, and the assessment of the outcome.^3^ In this meta-analysis, a NOS score of ≥6 stars was regarded as moderate- to high-quality, otherwise, as low-quality studies.^4^

**Statistical Analyses**

Summary RRs and 95% CIs for a 5 unit increment in BMI were using a random effects model. We calculated study-specific slopes (linear trends) and 95% CIs from the natural logs of the reported RRs and CIs across categories of adiposity measures adiposity by using the method of Greenland and Longnecker^5^. We performed the non-linear dose-response analysis by using the robust error meta-regression method described by *Xu et al*.^6^ This method is based on a “one-stage approach” which treating each study as a cluster of the whole sample and considering the within study correlations by clustered robust error. It requires known levels of adiposity and RRs with variance estimates for at least two quantitative exposure categories.^6^ For studies that did not set the lowest adiposity group as a reference, data were transformed using a method described by *Hamling et al*.^7^ which requires the number of cases and participants in each category. If these data could not be obtained from an article, the evidence was not pooled. If the median or mean adiposity was not provided and reported in ranges, we estimated the midpoint of each category by averaging the lower and upper boundaries of that category. If the highest or lowest category was open-ended, we assumed that the open-ended interval length was the same as the adjacent interval.^8^ To assess the heterogeneity of RRs across studies, the I*^2^* (95% CI) statistic was calculated with the following interpretation: low heterogeneity, defined as I*^2^* < 50%; moderate heterogeneity, defined as I*^2^* 50% to 75%; and high heterogeneity, defined as I*^2^* >75%.^9^ Moreover, subgroup analyses were not carried out when the numbers of included studies in outcomes were limited (≤5). A *P* value < 0.05 was considered statistically significant.

**SUPPLEMENTAL TABLES**

**Table S1: Search strategy**
**PubMed database**

| Search Terms | Search Options | Results |
| --- | --- | --- |
| #1 | Body mass index | 161830 |
| #2 | Obesity | 297876 |
| #3 | Overweight | 224992 |
| #4 | waist circumference | 29,607 |
| #5 | waist-to-hip ratio | 12,126 |
| #6 | Fat | 260345 |
| #7 | Atrial fibrillation | 72926 |
| #8 | Arrhythmia | 106668 |
| #9 | Cardiac surgery | 61919 |
| #10 | #1 OR #2 OR #3 OR #4 OR #5 OR #6 | 610950 |
| #11 | #7 OR #8 | 161385 |
| #12 | #10 AND #11 AND #12 | 133 |

**Embase database**

| Search Terms | Search Options | Results |
| --- | --- | --- |
| #1 | Body mass index | 409274 |
| #2 | Obesity | 519279 |
| #3 | Overweight | 94124 |
| #4 | Fat | 384874 |
| #5 | waist circumference | 31321 |
| #6 | waist-to-hip ratio | 14837 |
| #7 | Atrial fibrillation | 151344 |
| #8 | Arrhythmia | 185903 |
| #9 | Cardiac surgery | 106424 |
| #10 | #1 OR #2 OR #3 OR #4 OR #5 OR #6 | 1039942 |
| #11 | #7 OR #8 | 306848 |
| #12 | #10AND #11 AND #12 | 487 |

**Cochrane library**

| Search Terms | Search Options | Results |
| --- | --- | --- |
| #1 | Body mass index | 32707 |
| #2 | Obesity | 33603 |
| #3 | Overweight | 14018 |
| #4 | Fat | 29528 |
| #5 | waist circumference | 1382 |
| #6 | waist-to-hip ratio | 787 |
| #7 | Atrial fibrillation | 11307 |
| #8 | Arrhythmia | 8493 |
| #9 | Cardiac surgery | 8361 |
| #10 | #1 OR #2 OR #3 OR #4 OR #5 OR #6 | 76579 |
| #11 | #7 OR #8 | 17475 |
| #12 | #10 AND #11 AND #12 | 35 |

**Table S2:** PRISMA CHECKLIST

| **Section/topic** | **#** | **Checklist item** | **Reported on page #** |
| --- | --- | --- | --- |
| **TITLE** | | |  |
| Title | 1 | Identify the report as a systematic review, meta-analysis, or both. | 1 |
| **ABSTRACT** | | |  |
| Structured summary | 2 | Provide a structured summary including, as applicable: background; objectives; data sources; study eligibility criteria, participants, and interventions; study appraisal and synthesis methods; results; limitations; conclusions and implications of key findings; systematic review registration number. | 1-2 |
| **INTRODUCTION** | | |  |
| Rationale | 3 | Describe the rationale for the review in the context of what is already known. | 3 |
| Objectives | 4 | Provide an explicit statement of questions being addressed with reference to participants, interventions, comparisons, outcomes, and study design (PICOS). | 3-4 |
| **METHODS** | | |  |
| Protocol and registration | 5 | Indicate if a review protocol exists, if and where it can be accessed (e.g., Web address), and, if available, provide registration information including registration number. | 4 |
| Eligibility criteria | 6 | Specify study characteristics (e.g., PICOS, length of follow-up) and report characteristics (e.g., years considered, language, publication status) used as criteria for eligibility, giving rationale. | 4 |
| Information sources | 7 | Describe all information sources (e.g., databases with dates of coverage, contact with study authors to identify additional studies) in the search and date last searched. | 4 |
| Search | 8 | Present full electronic search strategy for at least one database, including any limits used, such that it could be repeated. | 4 |
| Study selection | 9 | State the process for selecting studies (i.e., screening, eligibility, included in systematic review, and, if applicable, included in the meta-analysis). | 4 |
| Data collection process | 10 | Describe method of data extraction from reports (e.g., piloted forms, independently, in duplicate) and any processes for obtaining and confirming data from investigators. | 4 |
| Data items | 11 | List and define all variables for which data were sought (e.g., PICOS, funding sources) and any assumptions and simplifications made. | 4-5 |
| Risk of bias in individual studies | 12 | Describe methods used for assessing risk of bias of individual studies (including specification of whether this was done at the study or outcome level), and how this information is to be used in any data synthesis. | 4-5 |
| Summary measures | 13 | State the principal summary measures (e.g., risk ratio, difference in means). | 5 |
| Synthesis of results | 14 | Describe the methods of handling data and combining results of studies, if done, including measures of consistency (e.g., I^2^) for each meta-analysis. | 5 |

Page 1 of 2

| **Section/topic** | **#** | **Checklist item** | **Reported on page #** |
| --- | --- | --- | --- |
| Risk of bias across studies | 15 | Specify any assessment of risk of bias that may affect the cumulative evidence (e.g., publication bias, selective reporting within studies). | 5 |
| Additional analyses | 16 | Describe methods of additional analyses (e.g., sensitivity or subgroup analyses, meta-regression), if done, indicating which were pre-specified. | 5 |
| **RESULTS** | | |  |
| Study selection | 17 | Give numbers of studies screened, assessed for eligibility, and included in the review, with reasons for exclusions at each stage, ideally with a flow diagram. | 5 |
| Study characteristics | 18 | For each study, present characteristics for which data were extracted (e.g., study size, PICOS, follow-up period) and provide the citations. | 5-6 |
| Risk of bias within studies | 19 | Present data on risk of bias of each study and, if available, any outcome level assessment (see item 12). | 6 |
| Results of individual studies | 20 | For all outcomes considered (benefits or harms), present, for each study: (a) simple summary data for each intervention group (b) effect estimates and confidence intervals, ideally with a forest plot. | 6 |
| Synthesis of results | 21 | Present results of each meta-analysis done, including confidence intervals and measures of consistency. | 6-7 |
| Risk of bias across studies | 22 | Present results of any assessment of risk of bias across studies (see Item 15). | 7 |
| Additional analysis | 23 | Give results of additional analyses, if done (e.g., sensitivity or subgroup analyses, meta-regression [see Item 16]). | 7 |
| **DISCUSSION** | | |  |
| Summary of evidence | 24 | Summarize the main findings including the strength of evidence for each main outcome; consider their relevance to key groups (e.g., healthcare providers, users, and policy makers). | 7 |
| Limitations | 25 | Discuss limitations at study and outcome level (e.g., risk of bias), and at review-level (e.g., incomplete retrieval of identified research, reporting bias). | 8-9 |
| Conclusions | 26 | Provide a general interpretation of the results in the context of other evidence, and implications for future research. | 10 |
| **FUNDING** | | |  |
| Funding | 27 | Describe sources of funding for the systematic review and other support (e.g., supply of data); role of funders for the systematic review. | 12 |

*From:* Moher D, Liberati A, Tetzlaff J, Altman DG, The PRISMA Group (2009). Preferred Reporting Items for Systematic Reviews and Meta-Analyses: The PRISMA Statement. PLoS Med 6(6): e1000097. doi:10.1371/journal.pmed1000097

For more information, visit: **www.prisma-statement.org**.

Page 2 of 2

**Table S3. Studies excluded (n=25) with reasons**

| **Studies not included** | **Reasons (According to PICOS)** |
| --- | --- |
| Phan, 2016[1] | Without target data set:This is a meta-analysis |
| Auer, 2005[2] | Not the target exposure: BMI information was insufficient. |
| Chua, 2015[3] | Not the target exposure: BMI information was insufficient. |
| Dandale, 2014[4] | Without target data set: Only providing RR without 95%CI. |
| Drossos, 2014[5] | Not the target exposure: Pericardial fat. |
| Kokkonen, 2005[6] | Not the target exposure: Postoperative hemodynamics and low postoperative serum triiodothyronine. |
| Massom, 2014[7] | Not the target exposure: Circulating cardiac biomarkers (NT-proBNP and hs-cTnT). |
| Mauermann, 2010[8] | Not the target exposure: Hemofiltration during cardiopulmonary bypass. |
| Nishi, 2012[9] | Not the target exposure: Only mentioned in the baseline characteristics of study patients. |
| O’Neal, 2013[10] | Not the target exposure and outcome: Study the relationship between postoperative atrial fibrillation and long-term survival. |
| Parsaee, 2014[11] | Not the target exposure: BMI information was insufficient. |
| Pillarisetti, 2014[12] | Not the target exposure and outcome: Study the relationship between early postoperative atrial fibrillation and late recurrence of AF. |
| Shirzad M, 2010[13] | Not the target exposure: BMI information was insufficient. |
| Weidinger, 2014[14] | Not the target exposure: BMI categories was insufficient. |
| B. Richter, 2006[15] | Not the target population: Patients referred for catheter ablation of AF. |
| Jidéus, 2000[16] | Without target data set: Not providing BMI categories. |
| Ducceschi, 1999[17] | Not the target exposure: BMI information was insufficient. |
| Ahlsson, 2009[18] | Not the target exposure: BMI information was insufficient. |
| Hravnak, 2002[19] | Not the target exposure: BMI information was insufficient. |
| Gürbüz, 2014[20] | Count data or univariate analysis |
| Cao, 2013[21] | Count data or univariate analysis |
| Kinoshita, 2011[22] | Count data or univariate analysis |
| Lopez-Delgado, 2015 | Count data or univariate analysis |
| Orhan, 2004[23] | Count data or univariate analysis |
| Wigfield, 2006[24] | Count data or univariate analysis |
| Yin, 2015[25] | Count data or univariate analysis |

**Table S4**. Quality assessment of included studies

| Author  (Publication Year) | Newcastle-Ottawa Scale | | | | | | | | | |  |
| --- | --- | --- | --- | --- | --- | --- | --- | --- | --- | --- | --- |
|  | Selection | | | Comparability | | | Outcome | | | Total | Average  score |
|  | a | b | c | d | e | f | g | h | i |  |  |
| Alam, 2011 | * | * | * |  | * | * | * |  |  | 6 | 7.6 |
| Ao, 2015 | * | * | * |  | * | * | * | * | * | 8 |  |
| Bramer, 2011 | * | * | * | * | * | * | * | * | * | 9 |  |
| Brandt, 2001 | * | * | * |  | * | * | * | * | * | 8 |  |
| Banach, 2007 | * | * | * |  | * | * | * | * | * | 8 |  |
| Engelman, 1999 | * | * | * | * | * | * |  |  |  | 6 |  |
| El-Chami, 2012 | * | * | * | * | * | * |  |  |  | 6 |  |
| Erbil N, 2013 | * | * | * | * | * | * | * | * | * | 9 |  |
| Efird, 2016 | * | * | * |  | * | * |  |  |  | 6 |  |
| Echahidi, 2014 | * | * | * | * | * | * | * | * | * | 9 |  |
| Gao, 2016 | * | * | * | * | * | * |  | * | * | 8 |  |
| Ghanta, 2017 |  |  | * | * | * | * | * |  |  | 5 |  |
| Girard, 2009 | * | * | * | * | * | * | * | * | * | 9 |  |
| Habib, 2005 | * | * | * |  | * | * |  | * | * | 7 |  |
| Hakala, 2002 | * | * | * | * | * | * | * | * | * | 9 |  |
| Ivanovic, 2014 | * | * | * | * | * | * | * | * | * | 9 |  |
| Kitahara, 2017 | * | * | * | * | * | * |  | * | * | 8 |  |
| Kuduvallia, 2002 | * | * | * | * | * | * | * | * | * | 9 |  |
| Moulton, 1996 | * | * | * |  | * | * |  | * | * | 7 |  |
| Melduni, 2011 | * | * | * | * | * | * | * | * | * | 9 |  |
| Omer, 2016 | * | * | * | * | * | * | * | * | * | 9 |  |
| Pan, 2006 | * | * | * |  | * | * |  | * | * | 7 |  |
| Perrier, 2016 | * | * | * | * |  |  | * | * | * | 7 |  |
| Reeves, 2003 | * | * | * |  | * | * | * |  | * | 7 |  |
| Stamou, 2011 | * | * | * |  | * | * |  | * | * | 7 |  |
| Sun, 2011 | * | * | * | * | * | * | * | * | * | 9 |  |
| Tosello, 2015 | * | * | * | * |  |  | * | * | * | 7 |  |
| Tadic M, 2011 | * | * | * | * | * | * | * | * | * | 9 |  |
| Yap, 2007 | * | * | * | * | * | * |  | * | * | 8 |  |
| Zacharias, 2005 | * | * | * | * | * | * | * |  |  | 7 |  |

a. Representativeness of the exposed cohort.
b. Selection of the non-exposed cohort.
c. Ascertainment of exposure.
d. Demonstration that outcome of interest was not present at start of study.
e. Comparability of cohorts on the basis of the design or analysis (adjusted for age).
f. Comparability of cohorts on the basis of the design or analysis (adjusted for any other factor).
g. Assessment of outcome.
h. Was follow-up long enough for outcomes to occur. (>5 days).
i. Adequacy of follow-up of cohorts.

**SUPPLEMENTAL FIGURES**

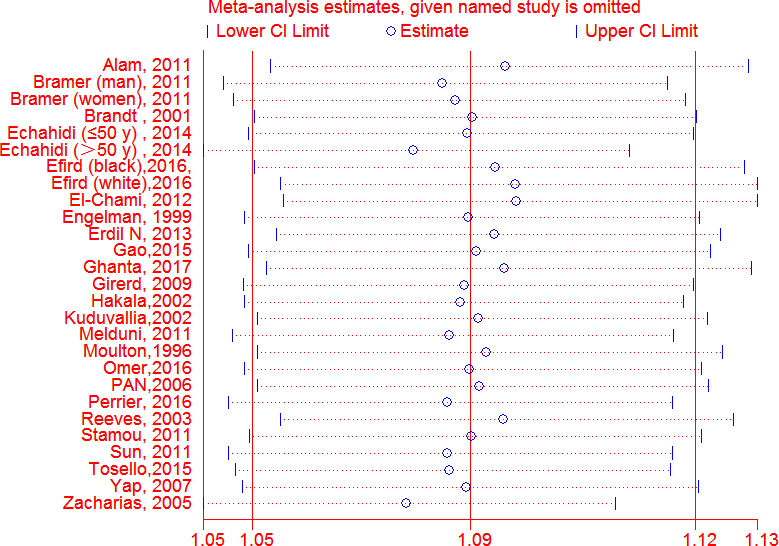

**Figure S1:** Sensitive analysis of BMI and POAF by omitting one study at each time, per 5 unit increased in BMI, dose-response analysis

Abbreviations: BMI = body mass index; SE=standard error; CI=confidence interval; POAF: atrial fibrillation after cardiac operation

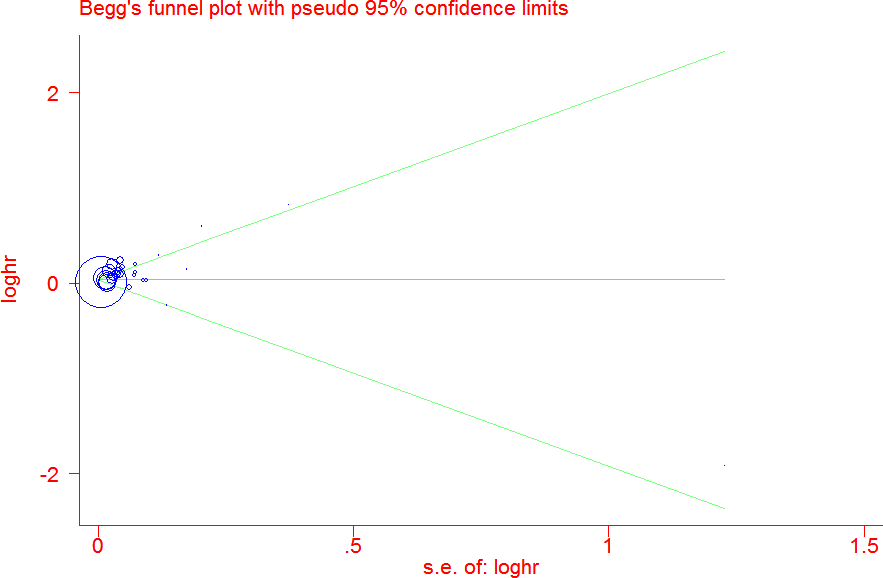

**Figure S2:** Publication bias analysis of the association between BMI and POAF, Begg’s test(P=0.73)

Abbreviations: BMI = body mass index; SE=standard error; POAF: atrial fibrillation after cardiac operation

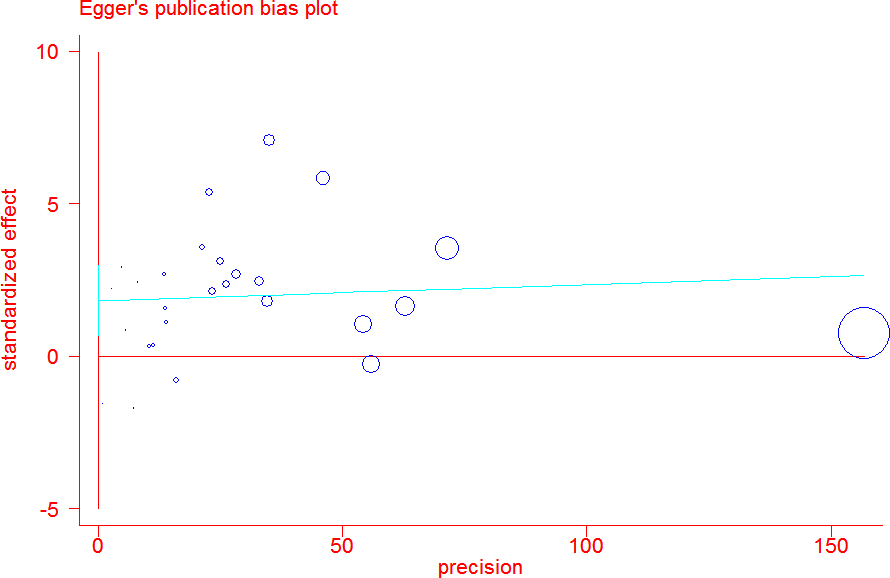

**Figure S3:** Publication bias analysis of the association between BMI and POAF, Egger’s test (P=0.42)

Abbreviations: BMI = body mass index; POAF: atrial fibrillation after cardiac operation

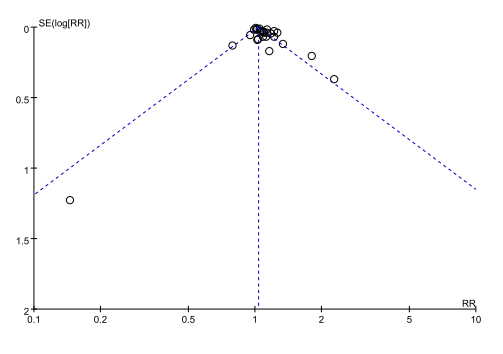

**Figure S4:** Publication bias analysis of the association between BMI and POAF, Funnel plot.

Abbreviations: BMI = body mass index; SE=standard error; POAF: atrial fibrillation after cardiac operation
